# Cutting a nerve of the hand alters the organisation of digit maps in primary somatosensory cortex

**DOI:** 10.1101/2024.11.26.24316490

**Authors:** Martin Weber, Andrew Marshall, Ronan Timircan, Francis McGlone, Raffaele Tucciarelli, Obi Onyekwelu, Louise Booth, Susan Francis, Michael Asghar, Edwin Jesudason, Vivien Lees, Kenneth F. Valyear

## Abstract

In non-human primates, forelimb nerve transection and repair alters the otherwise orderly and highly conserved organisation of the digit maps in primary somatosensory cortex (S1). These same reorganisational changes are presumed to occur in humans and to have meaningful implications for patient recovery. Using fMRI, we map digit responses in 21 patients with surgically repaired hand nerves and 30 controls. In S1 contralateral to the repaired hand, digit map arrangement is systematically altered, and response amplitudes are elevated. At the individual level, map organisation is more variable and less typical of controls, a pattern that, unexpectedly, is also evident in S1 of the uninjured hand. Our results show that hand nerve repair alters S1 organisation in humans, consistent with animal models. The functional relevance of these changes remains uncertain. We find no credible links between altered S1 maps and functional impairments.

## Introduction

Hand nerve injury constitutes a significant healthcare challenge. Incidence rates^1^ and economic costs^2^ are high, while patient recovery is typically incomplete^3,4^. Rehabilitation involves a complex interplay between peripheral and central factors^4,5^. Understanding how the brain changes and the clinical significance of those changes are major priorities.

When a nerve of the hand is cut and surgically repaired, nerve regeneration proceeds without topographical guidance^5–8^. Sprouting fibres innervate terminal receptors at different locations relative to the preinjury organisation, changing the structure of inputs from the hand to the central nervous system. In non-human primates, nerve transection and repair dramatically alters somatotopic organisation at multiple stations of the neuroaxis, including the fine-grained functional organisation of the digit maps in cortical areas 3b and 1^9–13^. Here, we evaluate whether similar brain changes occur in humans.

Primate somatosensory area 3b comprises a finely organised map of the hand^14,15^. Those neurons encoding nearby regions are grouped together forming spatially segregated functional maps. The relative spatial arrangement of these cortical maps is highly consistent across individuals. In the lateromedial direction, the maps for each digit are arranged in sequence, with interdigit map boundaries delineated by cell-poor septa^16^.

The functional maps of the hand are dramatically altered following nerve transection and repair in non-human primates^9–12^. Unlike the single continuous receptive fields observed normally, many cortical units now respond to multiple discontinuous locations on the hand, sometimes spanning multiple digits. Other cortical units exhibit normal single receptive fields, yet the typical spatial correspondence between the hand and cortex is altered. The digit maps are transformed from an orderly sequence that is highly conserved between individuals to a patchwork arrangement that is highly idiosyncratic. Touch of the same discrete patches of skin on the hand can also activate neurons that are non-adjacent within the cortical sheet, and some neurons exhibit large Pacinian-like receptive fields, not normally present in area 3b^14,15,17^.

The cortical changes described above depend on nerve regeneration in the periphery. Different changes are observed after nerve transection and ligation^18,19^, preventing nerve regeneration, as with amputation^20^. Purposeful crossing of peripheral nerves results in predictable changes in cortical topography^21^. Joining the proximal end of the ulnar nerve with the distal end of the median nerve, so that regenerating axons reinnervate the median-nerve-zone of the hand yet connect to the brain via the ulnar nerve, results in the emergence of cortical responses to touch of the median-nerve-zone of the hand within the ulnar-nerve-zone of cortex.

The current study examines whether nerve repair in humans alters the fine-grained organisation of the hand maps in primary somatosensory cortex (S1). The above evidence motivates two hypotheses. First, ***H_1_ decreased separability***, digit responses in S1 should be more difficult to distinguish in patients recovering from nerve repairs. The overall separability of interdigit responses should decrease. Second, ***H_2_ decreased typicality***, the relative spatial arrangement of the digit maps should change. Rather than the typical sequential ordering of D1-through-D5, nerve repair patients are expected to show an atypical patchwork arrangement, and the specific pattern of this arrangement is expected to show significant variation between individuals.

To evaluate these hypotheses, we use fMRI to quantify the pattern and extent of separability of interdigit responses in S1. While the precise position of individual digit response maps varies considerably between individuals, including the number and position of response ‘peaks’, the pattern of interdigit response separability is highly consistent^22,23^. Neighbouring digit responses are less separable, and the degree of separability between response maps generally declines as a function of their relative distance along the cortical sheet. This pattern reflects the frequency and co-occurrence of digit movements in everyday life ^22^. In nerve repair, misdirected nerve regeneration is expected to alter this pattern (Fig. 1).

**Figure 1.**
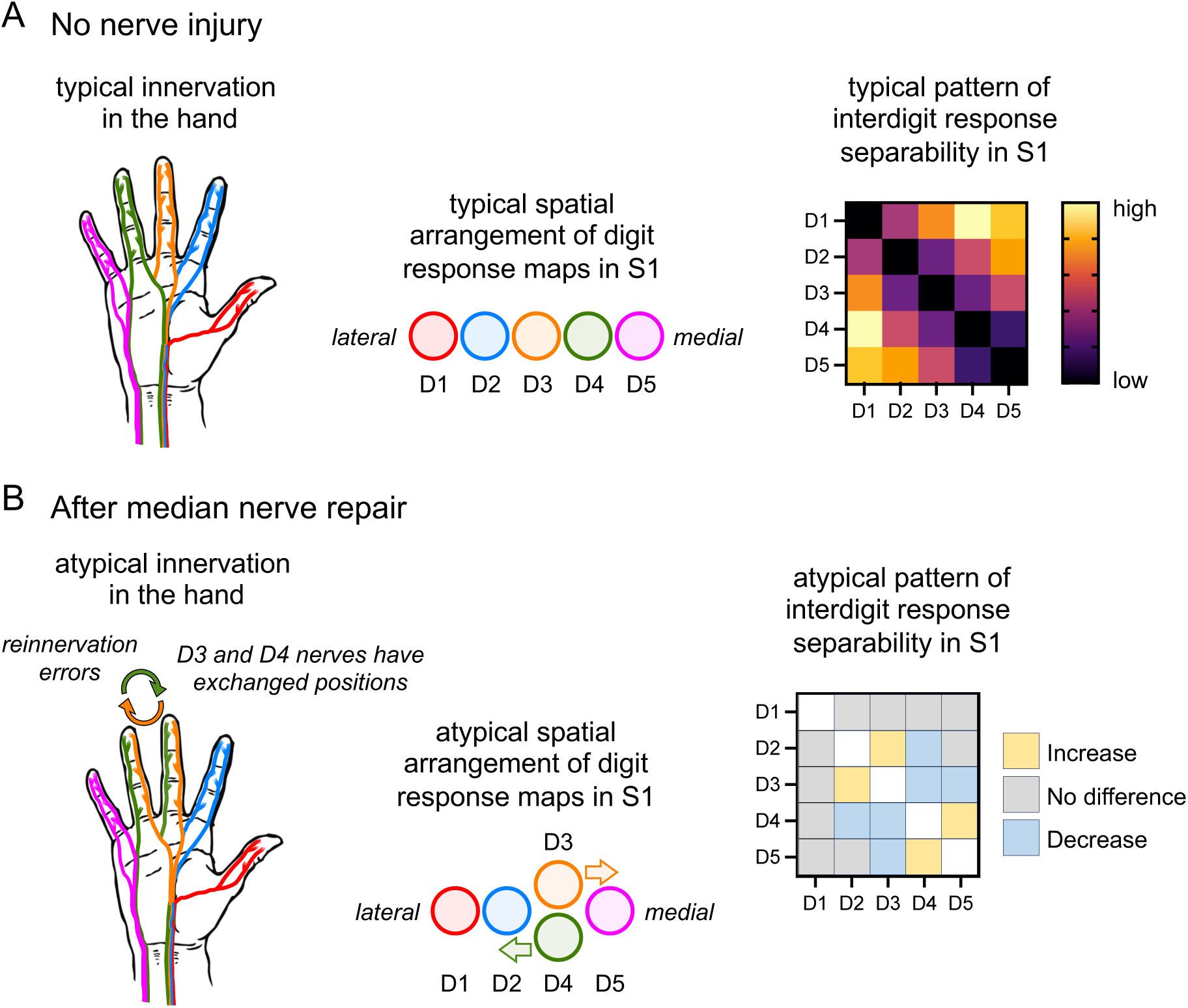
Reinnervation errors may alter the pattern of interdigit response separability in S1. **A:** The typical pattern of nerve innervation in the hand (left), the typical arrangement of digit maps in S1 (middle), and the typical pattern of interdigit response separability in S1 (right). The pattern of interdigit response separability is schematically estimated based on prior results^22^. Neighbouring digit maps are less separable, thumb responses are more distinct, and the separability of D1-vs-D4 is numerically larger than D1-vs-D5. **B:** After nerve transection and repair, regenerating fibres establish new connections without topographical guidance. The regrowth process is spontaneous and inherently random. Nerves may reroute to different digits or exchange between digits. Shown is a theoretical example where D3 and D4 nerves have exchanged positions after median nerve transection and repair (left). This would be expected to alter the relative spatial arrangement of digit response maps in S1 (middle) and the pattern of interdigit response separability (right). For simplicity, this is shown only for D3-D4 and their immediate neighbours; in principle, remapping could propagate further. The pattern would deviate from the otherwise highly stereotypical and conserved pattern defined in healthy controls.

Surprisingly few fMRI studies of hand nerve repair have been conducted—eight in total—and none have measured the detailed spatial arrangement of the digit maps in S1 ^24–31^. This work has typically involved small samples, focused mainly on median nerve injuries, and measured BOLD response amplitudes. Two consistent findings emerge. First, tactile stimulation of the repaired hand elicits elevated responses in contralateral S1. Second, elevated responses are also often observed ipsilaterally, in S1 contralateral to the uninjured hand. These ipsilateral effects are characterised by a loss of the typical deactivation (i.e., below-baseline responses) and have also been reported during stimulation of the uninjured hand—paralleling well-documented findings from upper-limb amputation^32^.

Increased fMRI responsivity after nerve repair is typically ascribed to cortical disinhibition^25^. These changes persist chronically, evident years after injury. One study found that these effects are associated with poor functional outcomes, and are absent in adults who sustained their injuries during childhood^28^. While these studies provide compelling evidence for functional brain changes in hand nerve repair, whether the stereotypical arrangement of the digit maps in S1 is altered remains unknown.

We also sought to determine whether brain changes in nerve repair have functional consequences, focusing primarily on touch localisation (locognosia). Locognosia errors in nerve repair may be explained by reinnervation errors^33,34^. Given that these reinnervation errors may also explain changes in the functional organisation of S1 (Fig. 1), it follows that changes in S1 may correlate with impaired locognosia.

## Results

We use 3T fMRI to map digit responses in 21 patients with hand nerve transections and 30 healthy controls. Vibrotactile stimulation is applied to distal digit pads (Supplementary Fig. S1), and S1 regions of interest are defined using a functional probabilistic atlas^35^ (Supplementary Fig. S2). Hypotheses regarding functional changes in nerve repair are specific to S1 contralateral to the repaired hand, *S1-patients-repaired*. Data from the uninjured hand (*S1-patients-uninjured*) and from healthy controls (*S1-controls*) serve as comparisons.

### Controls data reproduce the normative pattern of interdigit response separability

To evaluate our hypotheses, we quantify interdigit response separability in S1 using representational distances, computed as cross-validated squared Mahalanobis distances^36,37^. Critically, the normative pattern is known from previous research^22^ (Fig. 1A). Responses from topographically adjacent digit-maps, ‘1^st^ neighbours’, are less separable than those of 2^nd^, 3^rd^, and 4^th^ neighbours, and separability generally increases with increasing neighbourhood position.

Our results replicate previous findings. Representational distances (Fig. 2) reveal a systematic gradient of increasing interdigit response separability with increasing neighbourhood position. A significant main effect of neighbourhood position is confirmed (F(3, 87) = 106.1, p < 0.0001). Post-hoc comparisons reveal a stepwise increase in separability with increasing neighbourhood position (1^st^ > 2^nd^ > 3^rd^ > 4^th^, all t-values > 3.6, p-values < 0.005). The results validate our controls data for comparison against patients.

**Figure 2.**
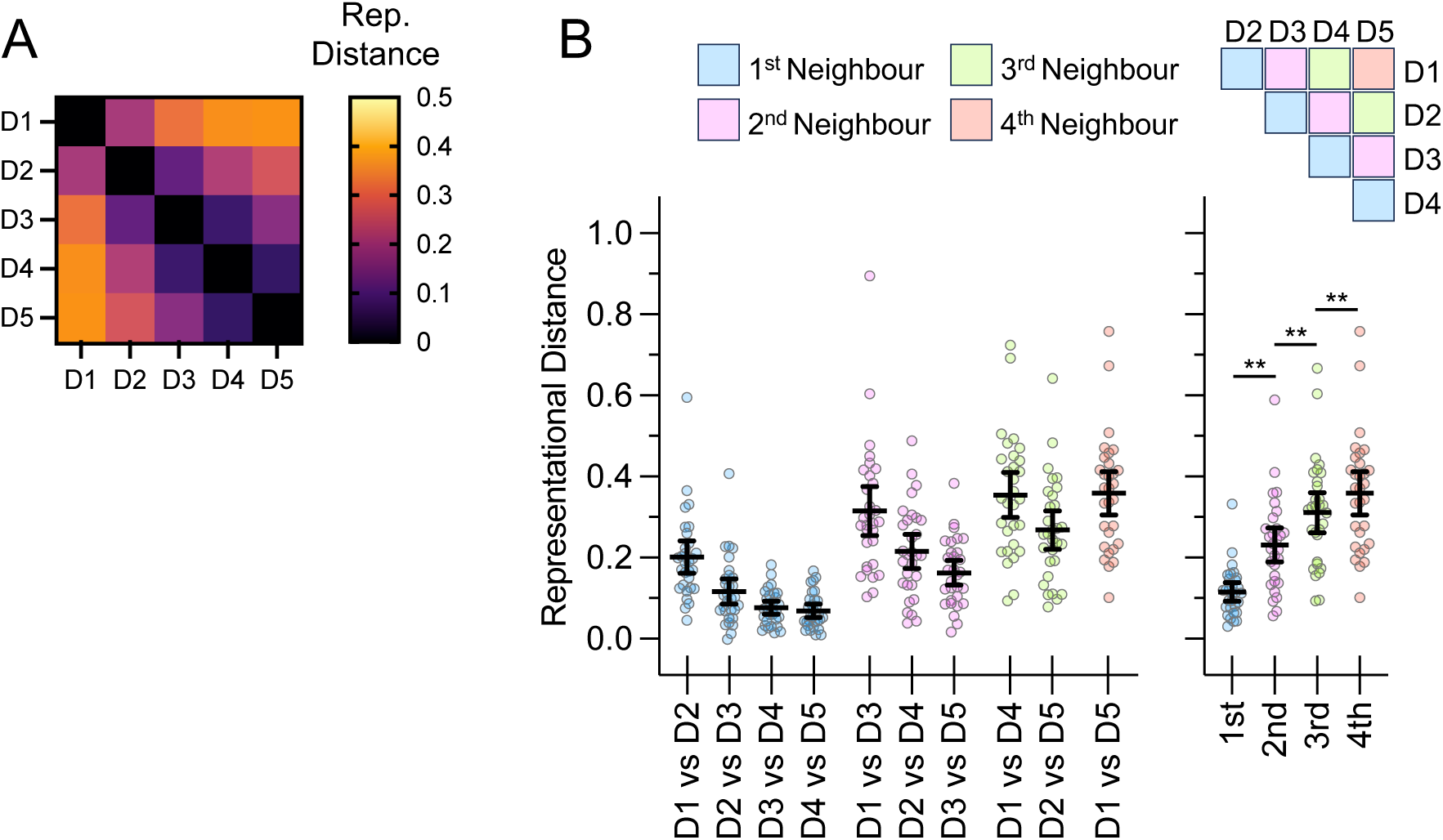
Interdigit response separability in healthy controls. **A:** Multivariate representational distances reflect the degree of dissimilarity between pairs of unthresholded digit responses. Greater values indicate more separable activation patterns. The heatmap shows the group mean representational distances across all possible (10) pairwise digit comparisons. **B:** Same data as in A, yet with individual-level results. Error bars are 95% confidence intervals around the group means. The results of each comparison are colour-coded according to neighbourhood position. The right-most plot shows the aggregate data. Note the stepwise increase in representational distances with increasing neighbourhood position. Asterisks indicate multiple-comparison-corrected significance (** p < 0.005; two-sided).

### Evaluating H_1_: Mean interdigit separability is preserved after nerve repair

Fig. 3A-B shows the group mean representational distances of patients and controls. The organisation is the same as in Fig. 2A-B; data from controls are repeated here for ease of comparison.

**Figure 3.**
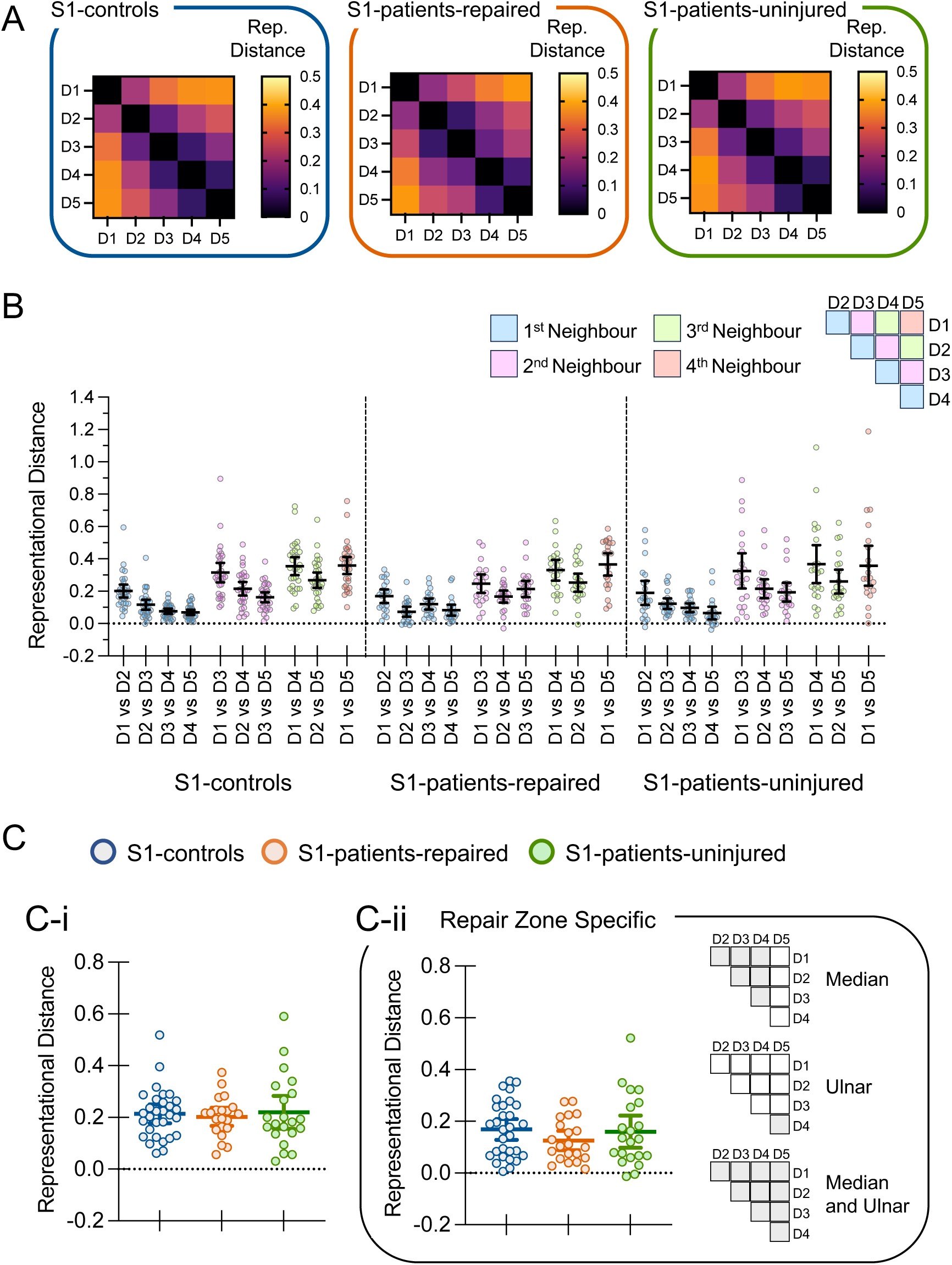
Evaluating H_1_: Mean interdigit separability. **A:** Group mean representational distances are shown as heatmaps for *S1-controls*, *S1-patients-repaired*, and *S1-patients-uninjured*. **B:** Same data as in A, yet with individual-level results. Error bars are 95% confidence intervals around the group means. Data are colour-coded according to neighbourhood position. **C-i:** Group representational distances averaged across all interdigit comparisons. **C-ii:** Group representational distances averaged across interdigit comparisons confined to the cortical zones of the repaired nerves. The icons illustrate how these zones differ between patient subgroups (grey = digit-pair included, white = digit pair not included).

Inconsistent with ***H_1_ decreased separability***, averaging representational distances across all interdigit comparisons reveals no consistent differences between patients and controls (Fig. 3C-i). Mean representational distances are similar between *S1-patients-repaired* and *S1-controls* (Mann-Whitney (U)=297, p=0.740; rank-biserial correlation (r_rb_)=-0.06; 95% confidence interval (95%-CI), [−0.05,0.04]; Bayes Factor (BF_10_)=0.31), and between *S1-patients-repaired* and *S1-patients-uninjured* (Wilcoxon W=31, p=0.609; r_rb_=0.13; 95%-CI, [−0.01,0.07]; BF_10_=0.24). As expected, no differences are observed between *S1-patients-uninjured* and *S1-controls* (U=296, p=0.726; r_rb_=-0.06; 95%-CI, [−0.07,0.06]; BF_10_=0.30).

Since monkey electrophysiological data indicate that changes in S1 are restricted to cortical zones of the repaired nerves^9–12^, we repeat these analyses using only those estimates of interdigit response overlap corresponding to maps within the nerve repair zones. Results are unchanged: *S1-patients-repaired* versus *S1-controls* (U=246, p=0.192; Holm–Bonferroni-corrected (H–B) p=0.575; r_rb_=-0.22; 95%-CI, [−0.11,0.02]; BF_10_=0.57); *S1-patients-repaired* versus *S1-patients-uninjured* (t(20)=1.14, p=0.266; H-B p=0.533; partial eta-squared (*η*^2^_p_)=0.06; 95%-CI, [−0.10,0.03]; BF_10_=0.41); *S1-patients-uninjured* versus *S1-controls* (U=277, p=0.476; r_rb_=-0.12; 95%-CI, [−0.09,0.05]; BF_10_=0.35).

### Evaluating H_2_: RDM structure is altered after nerve repair

Nerve repair alters the canonical digit-map structure in S1 (Fig. 4). A significant group × digit-pair interaction is observed between *S1-patients-repaired* and *S1-controls* (F(9,441)=4.12, p<0.001), demonstrating systematic alterations in the representational geometry—RDM structure—in patients compared to controls. The clearest departures are seen for interdigit comparisons involving D3 (Fig. 4D). These include reduced spacing for D1–D3, and increased spacing for D3–D4 and D3–D5 comparisons.

**Figure 4.**
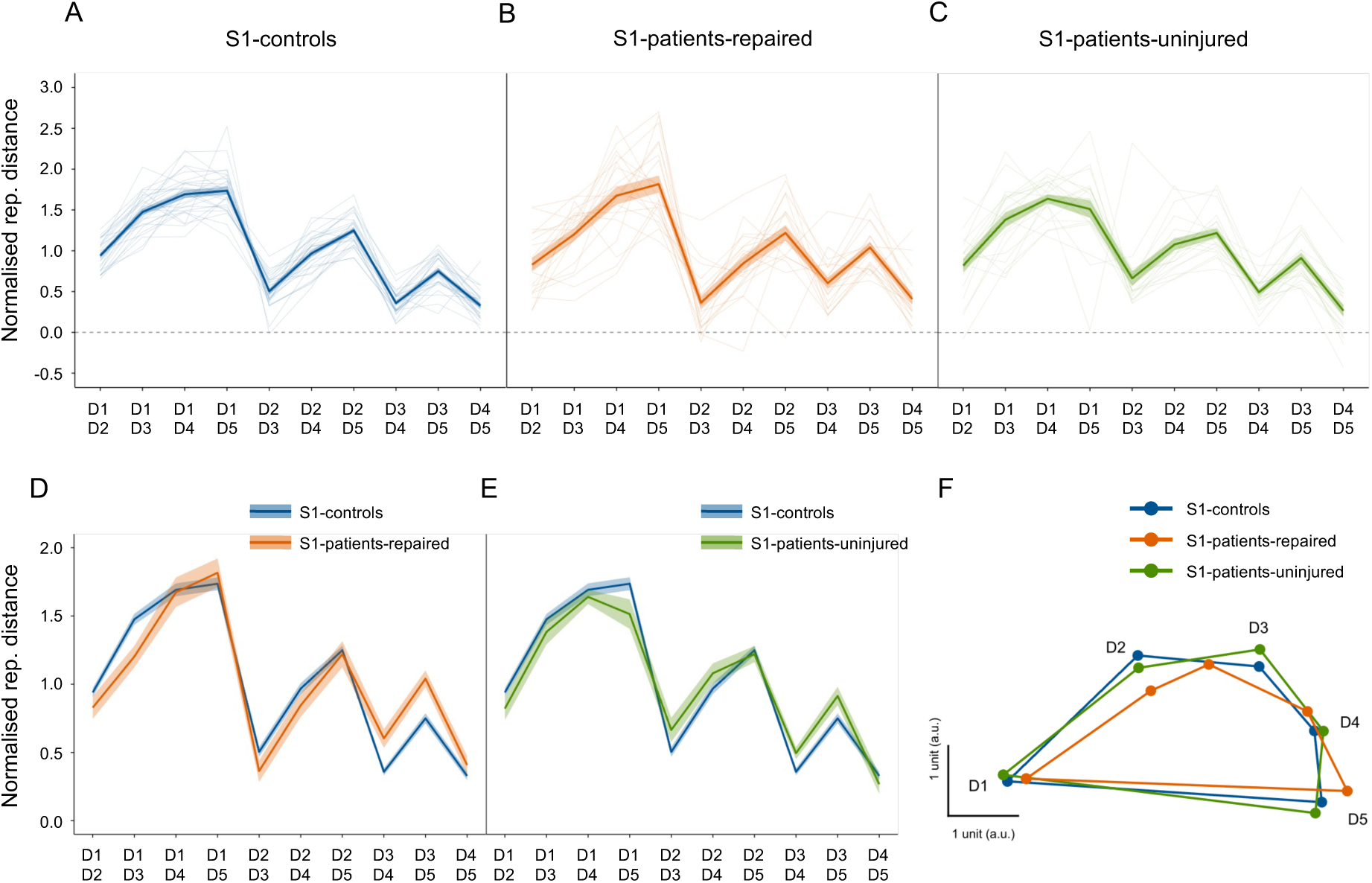
Evaluating H_2_: RDM structure. **A–C**: Normalised representational distances per interdigit comparison, shown as group means with shaded standard error bands: *S1-controls* (blue), *S1-patients-repaired* (orange), and S1-patients-uninjured (green). These are the same data shown in Fig. 3B, re-plotted after within-subject normalisation to highlight relative structure. Traces show individual participants. **D–E**: The group data from patients and controls are overlaid to facilitate comparisons (D: *S1-controls* vs. *S1-patients-repaired*; E: *S1-controls* vs. *S1-patients-uninjured*). **F**: Multidimensional scaling is used to project these data into a low-dimensional space to visualise the corresponding representational structure; the RDM structure in *S1-patients-repaired* is distorted.

Altered RDM structure is specific to the hemisphere contralateral to the repaired hand. A complementary mixed ANOVA comparing *S1-patients-uninjured* and *S1-controls* reveals no significant interaction (F(9,441)=0.32, p=0.967; BF_incl_=0.01), providing no evidence for changes in digit-map structure in S1 of the uninjured hand.

The results can be visualised with multidimensional scaling (Fig. 4F). The map in *S1-patients-repaired* is comparatively warped, especially at the position of D3. The results support ***H_2_ decreased typicality—***nerve repair transforms the otherwise highly conserved functional organisation of digit maps in S1.

Within-subject comparisons between hemispheres (*S1-patients-repaired* vs. *S1-patients-uninjured*) reveal inconclusive results. The ANOVA yields a borderline interaction effect (F(9,180)=1.85, p=0.062; BF_incl_=0.57), below statistical significance. Ultimately, the evidence is insufficient to support or refute a hemispheric difference within patients.

Main effects are consistent across all three ANOVAs. There are no significant main effects of group/hemisphere (all F’s < 0.30; all *p’s* > 0.600), consistent with the above results evaluating H_1_. Digit-pair showed significant main effects in all three ANOVAs (all F’s > 39.75; all *p’s* < 0.0001).

### Evaluating H_2_: RDM typicality is decreased after nerve repair

While the ANOVA tests for systematic group-level changes, typicality tests for individual-level changes, providing a measure of agreement between an individual’s interdigit response pattern and the normative pattern defined by healthy controls. High positive typicality indicates close agreement.

Fig. 5A provides examples of individual-level RDM typicality. Some patients closely resemble the normative pattern, while others do not. ID63 (isolated median repair) and ID26 (repairs to both nerves) show atypical patterns, with qualitative inspection of their heatmaps revealing clear differences from controls.

**Figure 5.**
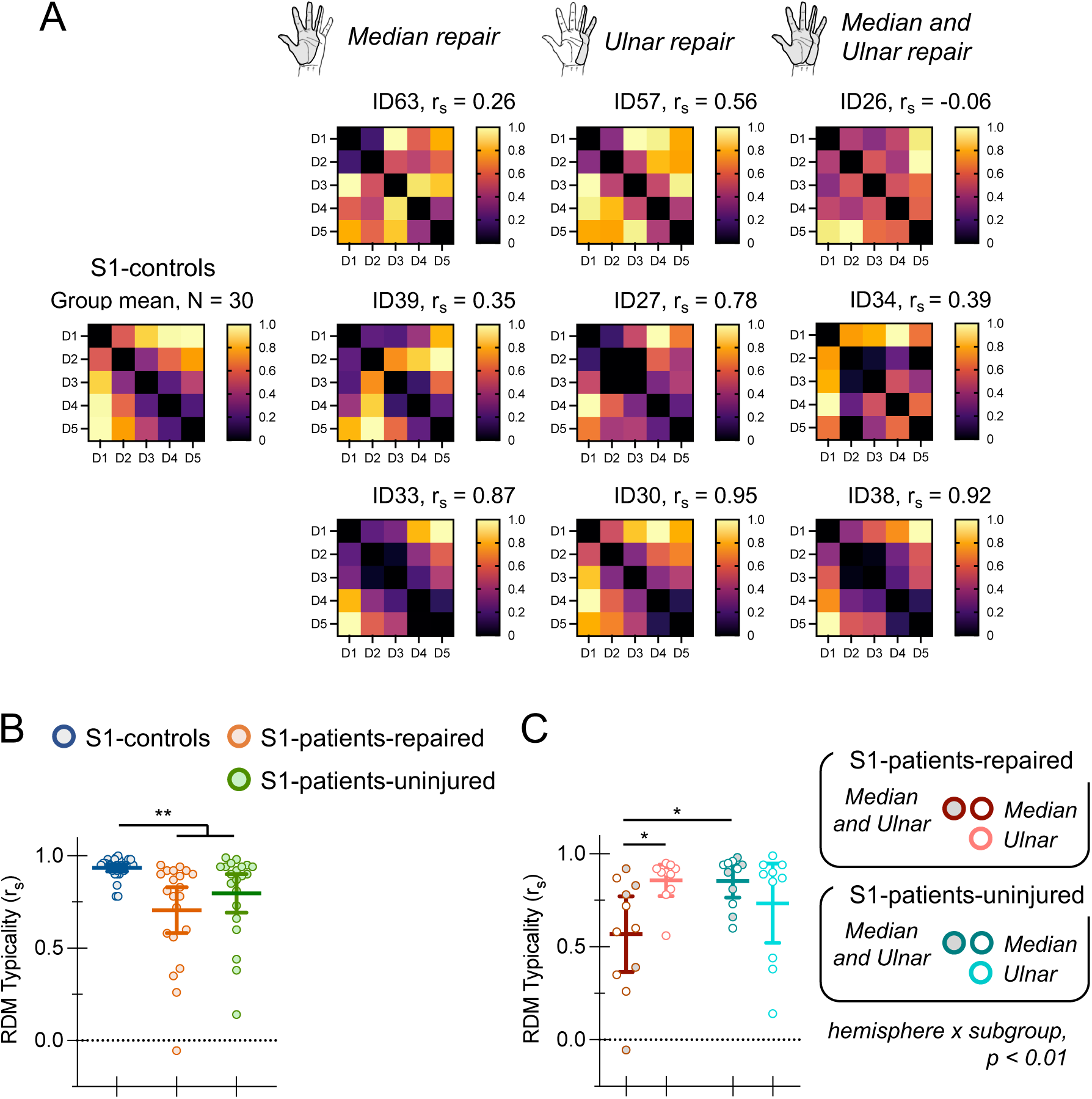
Evaluating H_2_: RDM typicality. **A:** Examples of individual RDM typicality in S1 of the repaired hand (*S1-patients-repaired)*. For each repair type (median, ulnar, both), the RDM heatmaps of patients with the lowest, second-lowest, and highest typicality values are shown. The *S1-controls* group-mean RDM heatmap is provided for reference. Representational distances are range normalised. **B:** Group RDM typicality for *S1-controls*, *S1-patients-repaired*, and *S1-patients-uninjured*; both hemispheres in patients show reduced typicality relative to controls. **C:** RDM typicality by patient subgroup and hemisphere. In *S1-patients-repaired*, typicality is lower after median or both nerve repairs than after ulnar repairs; no subgroup effect is observed on the uninjured side (*S1-patients-uninjured)*. Error bars (B, C) are 95% confidence intervals. Asterisks indicate multiple-comparison-corrected significance (* p < 0.05; ** p < 0.01; two-sided).

Quantitative group comparisons support ***H_2_ decreased typicality*** (Fig. 5B). Nerve repair significantly alters the otherwise stereotypical pattern of interdigit responses observed in healthy controls. RDM typicality is decreased in *S1-patients-repaired* compared to *S1-controls* (U=70.5, p < 0.001; H-B p < 0.001, r_rb_ =-0.78; 95%-CI, [0.06,0.24]). Unexpectedly, typicality is also decreased for *S1-patients-uninjured* compared to *S1-controls* (U=162.5, p=0.003; H-B p=0.006; r_rb_=-0.48; 95%-CI, [0.01,0.11]), and *S1-patients-repaired* does not differ from *S1-patients-uninjured* (t(20)=1.18, p=0.251; η^2^_p_=0.07; 95%-CI, [−0.25,0.07]; BF_10_=0.42). In other words, altered S1 maps are observed bilaterally.

### Decreased typicality on the repaired side depends on the nerve(s) repaired

Because primate models show that S1 remodelling is restricted to the nerve-repair zone, one might expect isolated ulnar nerve repairs to produce subtler changes. To investigate this, we performed a patient subgroup analysis of RDM typicality separating ulnar nerve repairs from median and both repairs.

Fig. 5C shows these results. In *S1-patients-repaired*, typicality depends on the nerves affected. Patients with repairs to the median or ‘both’ nerves have significantly lower typicality compared to patients with isolated ulnar repairs. These effects are specific to S1 of the repaired hand. In *S1-patients-uninjured*, typicality does not depend on patient subgroup. A mixed ANOVA reveals a significant interaction (F(1,19)=10.27, p=0.005; η^2^_p_=0.36), and no significant main effects (hemisphere: F(1,19)=1.63, p=0.217; η^2^_p_=0.08; BF_Incl_=3.09; subgroup: F(1,19)=1.19, p=0.289; η^2^_p_=0.06; BF_Incl_=3.36). Post-hoc comparisons confirm patient subgroup differences in *S1-patients-repaired* (t(19)=2.83, p=0.011; H-B p=0.021; η^2^=0.30; 95%-CI, [0.08, 0.50]) but not in *S1-patients-uninjured* (U=41.5, p=0.357; r_rb_=0.25; 95%-CI, [−0.08, 0.37]; BF_10_=0.53).

The findings indicate that the degree of cortical reorganisation in S1 of the repaired hand depends on which nerve(s) were repaired. Patients with repairs to the median or both median and ulnar nerves exhibit lower typicality than those with isolated ulnar repairs, consistent with more extensive reorganisation when a larger portion of the hand—and thus S1—is involved. This aligns with primate evidence indicating that S1 remodelling is restricted to the nerve-repair zone.

These same findings can be described differently. Holding patient subgroup constant, hemispheric differences emerge only for median/both repairs: typicality is lower in *S1-patients-repaired* compared to *S1-patients-uninjured* (t(19)=2.97, p=0.014; H–B p=0.028; η^2^_p_=0.47; 95%-CI, [0.07, 0.50]; Fig. 5C), with no hemispheric difference for ulnar-only repairs (W=19, p=0.361; r_rb_=0.35; 95%-CI, [−0.05,0.54]; BF_10_=0.53; Fig. 5C). Moreover, typicality in *S1-patients-repaired* does not correlate with typicality in *S1-patients-uninjured* (r_s_(19)=0.05, p=0.839; 95%-CI, [−0.40,0.48]; BF_10_=0.30; Supplementary Fig. S3). Together, these results are inconsistent with a mirrored interhemispheric remapping account wherein the alterations in S1 of the uninjured hand directly reflect—mirror—the alterations in S1 of the repaired hand (cf. animal deafferentation models^38,39^).

### Decreased typicality is not explained by decreased measurement reliability

Reduced typicality of the patient’s uninjured side (S1-*patients-uninjured*) is unexpected and raises an important interpretive challenge. Might decreased RDM typicality in patients reflect noisier measurements, rather than genuine alterations in the functional organisation of S1 digit maps?

Perhaps our patient data are more variable across imaging runs, giving rise to unreliable estimates of the true representational geometry of the digit maps, and thus spuriously lower RDM typicality. Our RDMs reflect cross-validated squared Mahalanobis distances, combining run-wise cross-validation with voxel-wise noise normalisation to minimise the influence of noise^37^. Cross-validation ensures that distances are computed using independent data folds, such that noise shared across conditions or runs does not inflate similarity estimates, while pre-whitening down-weights noisy or correlated voxels, improving sensitivity to consistent patterns of neural activity. Nonetheless, it remains possible that representational geometry in patients is less stable across imaging runs, leading to greater variability in the RDM structure and consequently to lower typicality.

To evaluate this possibility, we tested the within-subject reliability of interdigit representational geometry across imaging runs. For each participant and each cross-validation fold, we computed the Pearson correlation coefficient between the RDM from the held-out run (test set) and the average RDM from the remaining four runs (train set). These fold-wise correlations were then averaged to yield a single reliability score per subject per hemisphere.

This procedure provides a conservative test of internal consistency. If patients show lower reliability scores than controls, reduced typicality may reflect unstable representations rather than meaningful reorganisation. Conversely, if within-subject reliability is comparable across groups, this would argue against noise or instability as an explanatory factor in the observed group differences in RDM typicality.

We find no evidence that RDM structure is less reliable in patients (Supplementary Fig. S4). No evidence is found for differences between *S1-patients-repaired* and *S1-controls* (*t*(49)=0.35, p=0.727; η^2^=0.003; 95%-CI, [−0.07,0.09]; BF_10_=0.30), *S1-patients-uninjured* and *S1-controls* (*t*(49)=0.46, *p*=0.648; η^2^=0.004; 95%-CI, [−0.06,0.09]; BF_10_=0.31), or *S1-patients-repaired* and *S1-patients-uninjured* (*t*(20)=0.07, *p*=0.945; η^2^ =0.0002; 95%-CI, [−0.08,0.08]; BF_10_=0.23). Reliability values are modest overall, as expected for single-run comparisons, but critically, they do not differ between patients and controls. These findings argue against the possibility that reduced typicality in patients is driven by increased measurement noise or instability of representational geometry across imaging runs.

### BOLD response amplitudes are elevated on the repaired side

Although not a direct test of our hypotheses, we analyse S1 response amplitudes—percent BOLD signal change (%-BSC) values—for two reasons. First, weaker perceived touch on the repaired hand might evoke diminished BOLD response amplitudes in S1, and although representational geometry—i.e., the relative structure of the RDMs—is largely stable across a wide range of BOLD activity levels, lower signal can compress representational distances^40^. Second, in contrast to this concern, previous fMRI studies of hand nerve repair consistently report increased BOLD activity in S1^24,25,27,28,30,31^. Tactile stimulation of the repaired hand elicits higher-than-typical %-BSC values, often observed bilaterally, in both contra- and ipsilateral S1.

Our results reveal increased BOLD response levels in patients compared with controls, specific to the repaired hand and the contralateral hemisphere (Fig. 6A). In *S1-patients-repaired*, %-BSC values are significantly higher than in both *S1-controls* (*t*(49)=2.47, p=0.017; H-B p=0.033; η^2^=0.11; 95%-CI, [0.03,0.27]) and *S1-patients-uninjured* (*t*(20)=3.65, p=0.002; H-B p=0.005; η^2^=0.40; 95%-CI, [0.07,0.26]). Stimulation of the uninjured hand results in similar BOLD response amplitudes as controls: we find no evidence for differences between *S1-patients-uninjured* and *S1-controls* (*t*(49)=0.45, p=0.657; η^2^=0.004; 95%-CI, [−0.12,0.07]; BF_10_=0.31).

**Fig. 6.**
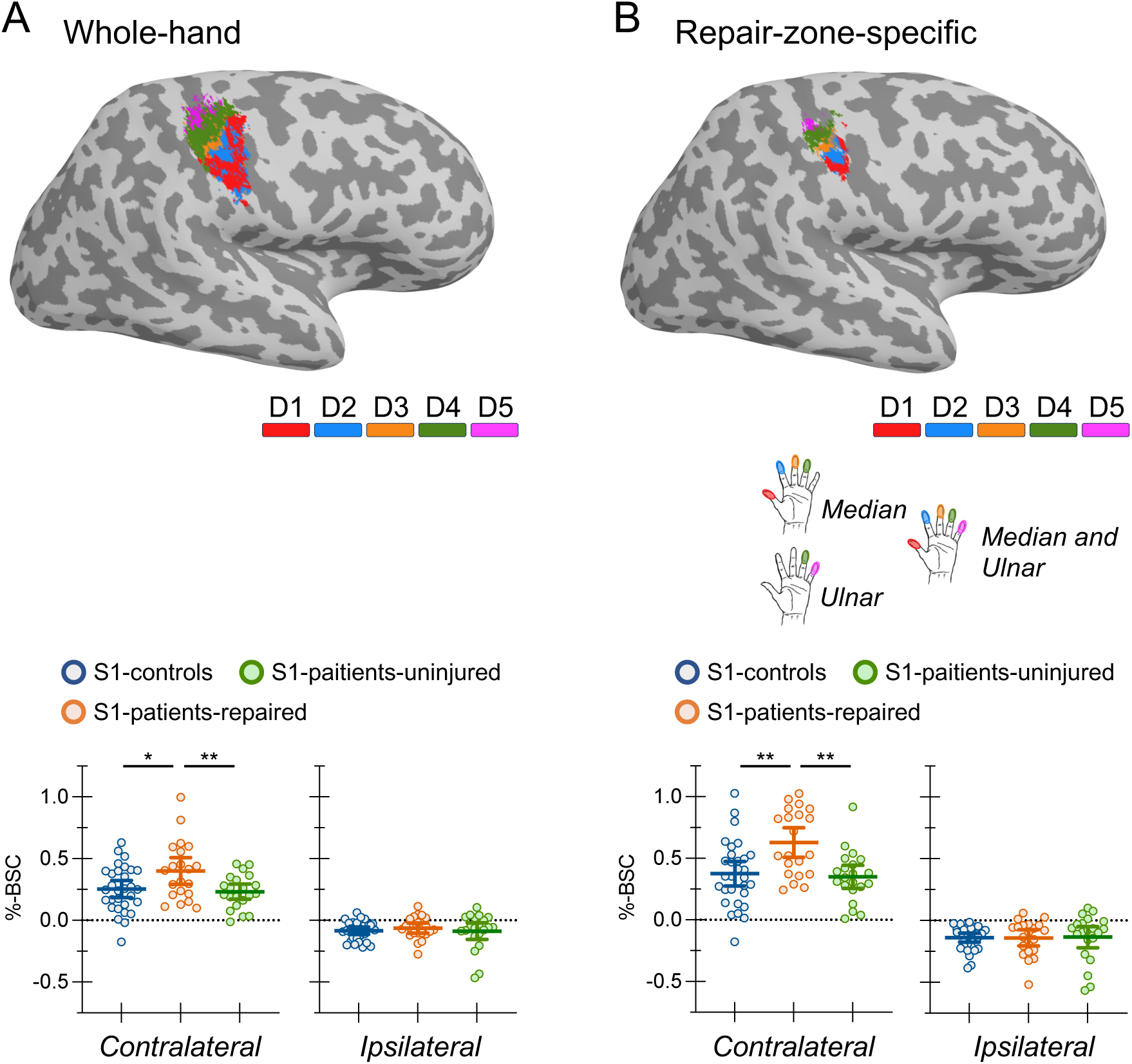
Percent BOLD change (%-BSC) **A:** Whole-hand analyses. Top: cortical surface maps showing atlas-defined digit-specific ROIs projected onto an individual participant’s surface. Bottom: group mean %-BSC values corresponding to stimulation of the contralateral (left) and ipsilateral (right) hand for *S1-controls*, *S1-patients-repaired*, and *S1-patients-uninjured*. Contralateral BOLD response amplitudes are elevated for *S1-patients-repaired.* Ipsilateral responses show no group differences; patients and controls show the expected below-baseline response levels. **B:** Repair-zone-specific analyses. Same layout as A; here, ROIs are defined using higher thresholds and restricted to the cortical repair zone, based on patient repair type (median, ulnar, or both), with extracted responses taken only from repaired digits (homotopic digits on the uninjured side and in controls; see Methods). Results replicate those of the whole-hand analyses. Error bars are 95% confidence intervals. Asterisks indicate multiple-comparison-corrected significance (* p < 0.05; ** p < 0.01; two-sided).

Conversely, response amplitudes in ipsilateral S1 do not differ between groups (Fig. 6A). No differences are observed between *S1-patients-repaired* and *S1-controls* (*t*(49)=0.93, p=0.359; η^2^=0.02; 95%-CI, [−0.03,0.07]; BF_10_=0.40), *S1-patients-uninjured* and *S1-controls* (U=272, p=0.419; r_rb_=-0.14; 95%-CI, [−0.04,0.08]; BF_10_=0.34), or *S1-patients-repaired* and *S1-patients-uninjured* (W=11, p=0.865; r_rb_=0.05; 95%-CI, [−0.08,0.10]; BF_10_=0.26). Patients and controls show the typical below-baseline BOLD signal effects on the ipsilateral side. This result contrasts with previous reports of elevated ipsilateral activity in nerve repair.

Whole-hand ROIs could dilute effects specific to reinnervated digits. We therefore repeated these analyses using more selective thresholds to define digit-specific ROIs confined to the cortical repair zone, extracting responses only from stimulation of the corresponding digits.

The results replicate those of the whole-hand analyses (Fig. 6B): contralateral %-BSC remains elevated for *S1-patients-repaired* relative to *S1-controls* (t(49)=3.36, p=0.002; H–B p=0.003; η^2^=0.19; 95%-CI, [0.10,0.41]) and *S1-patients-uninjured* (t(20)=5.00, p<0.001; H–B p<0.001; η^2^= 0.56; 95%-CI, [0.16,0.40]), with no difference between *S1-patients-uninjured* and *S1-controls* (t(49)=0.36, p=0.717; η^2^=0.002; 95%-CI, [−0.16,0.11]; BF_10_=0.30). Ipsilateral results are also unchanged (Fig. 6B): *S1-patients-repaired* vs *S1-controls* (*t*(49)=0.035, *p*=0.972; η^2^<0.0001; 95%-CI, [−0.06,0.06]; BF_10_=0.28); *S1-patients-uninjured* vs *S1-controls* (*t*(49)=0.10, *p*=0.919; η^2^< 0.001; 95%-CI, [−0.08,0.09]; BF_10_=0.29); *S1-patients-repaired* vs *S1-patients-uninjured* (*t*(20)=0.14, *p*=0.893; η^2^<0.001; 95%-CI, [−0.09,0.08]; BF_10_=0.23).

Altogether, these results provide two insights. First, group differences in RDM structure (Fig. 4) and RDM typicality (Fig. 5) are not confounded by reduced BOLD effects in patients due to impaired touch sensitivity (see also Supplementary Fig. S5 and S6). Second, elevated contralateral S1 responses to stimulation of the repaired hand provide additional evidence for cortical plasticity following peripheral nerve repair, consistent with previous reports.

### Inconclusive evidence for a relationship between altered S1 maps and impaired locognosia

Next, we evaluate whether brain changes after nerve repair have functional consequences for touch localisation (locognosia). Because reinnervation errors can produce locognosia errors^33,34^ and, in principle, distort S1 organisation (Fig. 1), we predicted that altered maps would correlate with impaired locognosia.

Our fMRI results provide two markers of altered maps—changes in RDM structure (Fig. 4) and reduced RDM typicality (Fig. 5). Of these, only RDM typicality provides an individual-level measure suitable for correlational tests.

Nineteen patients completed locognosia testing using the Digital Photograph Method (see Methods and Supplementary Fig. S7), with detailed results from eighteen patients (and thirty-three controls) reported previously^41^.

Absolute error measures the difference between stimulated and perceived locations of touch, excluding misreferrals—errors made between digits, or from a digit to the palm. Lower absolute error indicates better performance. A single patient’s data is shown in Fig. 7A, for illustration.

**Figure 7.**
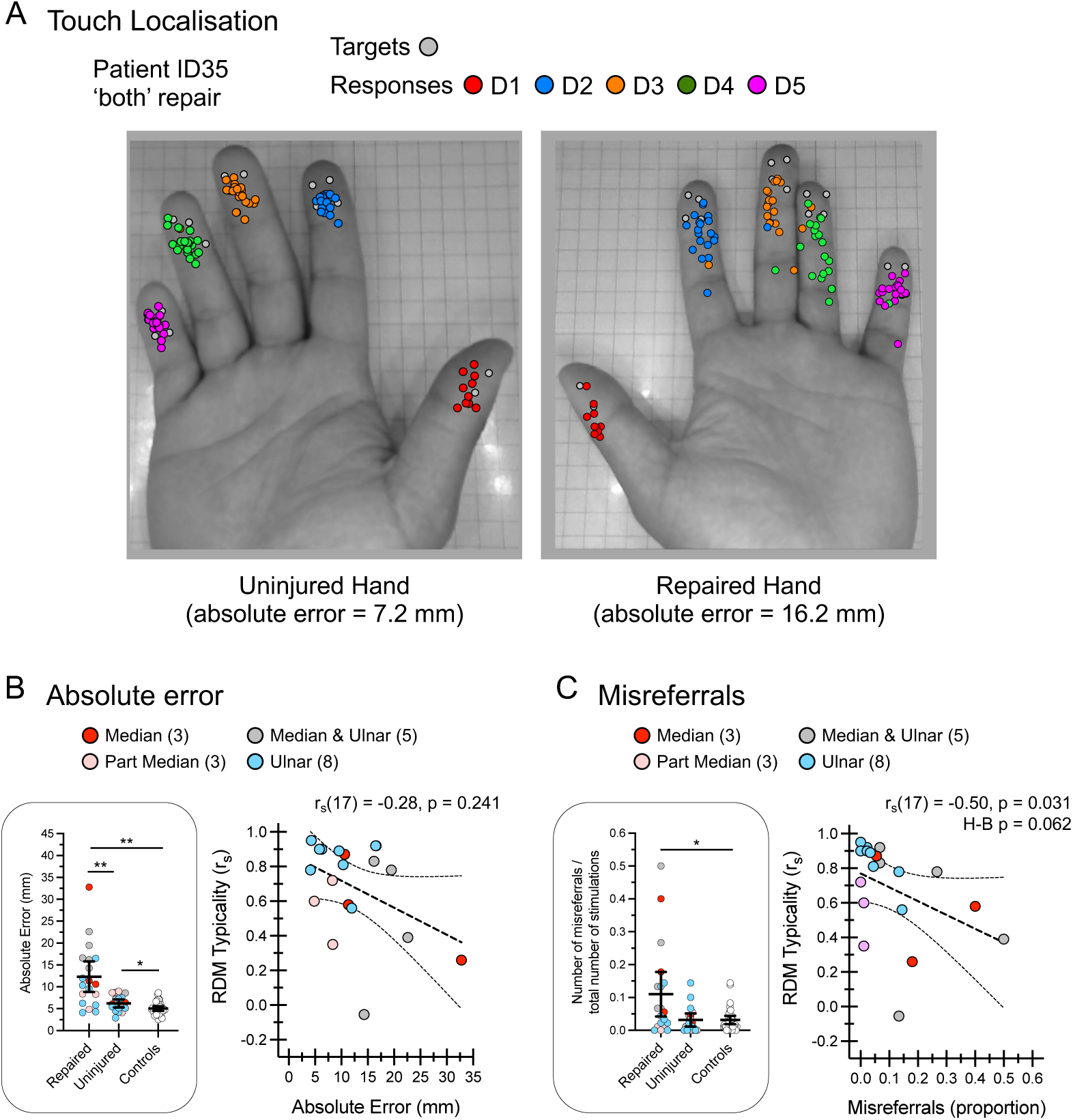
Inconclusive evidence for a relationship between altered S1 maps and impaired locognosia. **A:** Locognosia results from a single patient with combined median and ulnar repairs. Targets (grey circles) and responses (coloured circles) are overlaid on the photographs of the hand. The colours indicate which digit was touched (see inset key). The repaired hand shows a larger spread of responses (higher absolute error). Absolute error excludes misreferrals. **B:** Absolute error. *Left*: group mean absolute error for the repaired hand (N=19), uninjured hand (N=18), and controls (N=33). Statistical results: repaired vs controls (U=81, p<0.001; H–B p<0.001; r_rb_=0.74; 95%-CI, [3.44, 9.17]); repaired vs uninjured (W=149, p<0.001; H–B p<0.001; r_rb_=0.87; 95%-CI, [0.72, 9.02]); uninjured vs controls (t(49)=2.54, p=0.014; *η*^2^=0.12; 95%-CI, [0.25, 2.13]). *Right*: Absolute error (repaired hand) versus RDM typicality for *S1-patients-repaired*. No evidence for a relationship is found. **C:** Misreferrals. *Left*: group mean misreferrals expressed as a proportion of total trials (90) for the repaired hand, uninjured hand, and controls. Statistical results (arcsine-transformed): repaired vs controls (U=182, p=0.011; H–B p=0.034; r_rb_=0.42; 95%-CI, [0.03, 0.21]); repaired vs uninjured (t(17)=2.27, p=0.037; H–B p=0.073; *η*^2^=0.23; 95%-CI, [0.01, 0.26]; BF_10_=1.84); uninjured vs controls (U=291, p=0.910; r_rb_=0.02; 95%-CI, [−0.05, 0.08]; BF_10_=0.30). *Right*: Misreferrals (repaired hand) versus RDM typicality for *S1-patients-repaired*; the trend does not survive Holm-Bonferroni-correction for multiple comparisons. Error bars are 95% confidence intervals. Asterisks indicate multiple-comparison-corrected significance (* p<0.05; ** p<0.001; two-sided). ‘Part median’ indicates patients with incomplete median nerve transection injuries

We find no reliable evidence for a relationship between absolute error and RDM typicality (r_s_(17)=-0.28, p=0.241; 95%-CI, [−0.66,0.21]; BF_10_=0.84) (Fig. 7B). Patients who make larger within-digit errors in localising punctate, suprathreshold touch stimulation on their repaired hand do not, on average, show lower typicality.

Misreferrals are between-digit (or digit-to-palm) errors. Most patients make few misreferrals, and only four patients exceed the range observed in healthy controls (Fig. 7C). This leaves little between-participant variance and thus low power for group-level correlations.

Nevertheless, misreferrals tend to increase as typicality decreases (r_s_(17)=−0.50, p=0.031; H–B p=0.062; 95%-CI, [−0.78,-0.03]; BF_10_=3.47; Fig. 7C). This result does not survive correction for multiple comparisons and is therefore inconclusive. Influence checks indicate the trend is not reducible to a single participant. Given the potential clinical significance of establishing a relationship between altered S1 maps and functional impairments, well-powered follow-up studies involving paradigms optimised for the measurement of between-digit errors are needed.

Descriptive results from two patients with unusual frequencies and patterns of misreferrals are useful to visualise the potential links between misreferrals, reinnervation errors, and altered digit maps. We offer these results for conceptual purposes.

Patient ID21 consistently reports touch of the index finger as being felt on the middle finger, and sometimes vice versa (Fig. 8A). In other words, the index and middle fingers are often confused; a pattern not observed in healthy controls^41^. In principle, this could result from an exchange between digital nerves (Fig. 8B). Such reinnervation errors would predict specific alterations in the arrangement of the digit maps in S1, and thus the pattern of interdigit response separability. Fig. 8C compares the patient’s pattern of representational distances against the normative pattern in controls. Some differences align with predictions (e.g., decreased D2-D3 separability), while others do not (e.g., increased D1-D5 separability). Patient ID32 also exhibits a pattern of misreferrals that could reflect reinnervation errors between digits (Supplementary Fig. S8).

**Figure 8.**
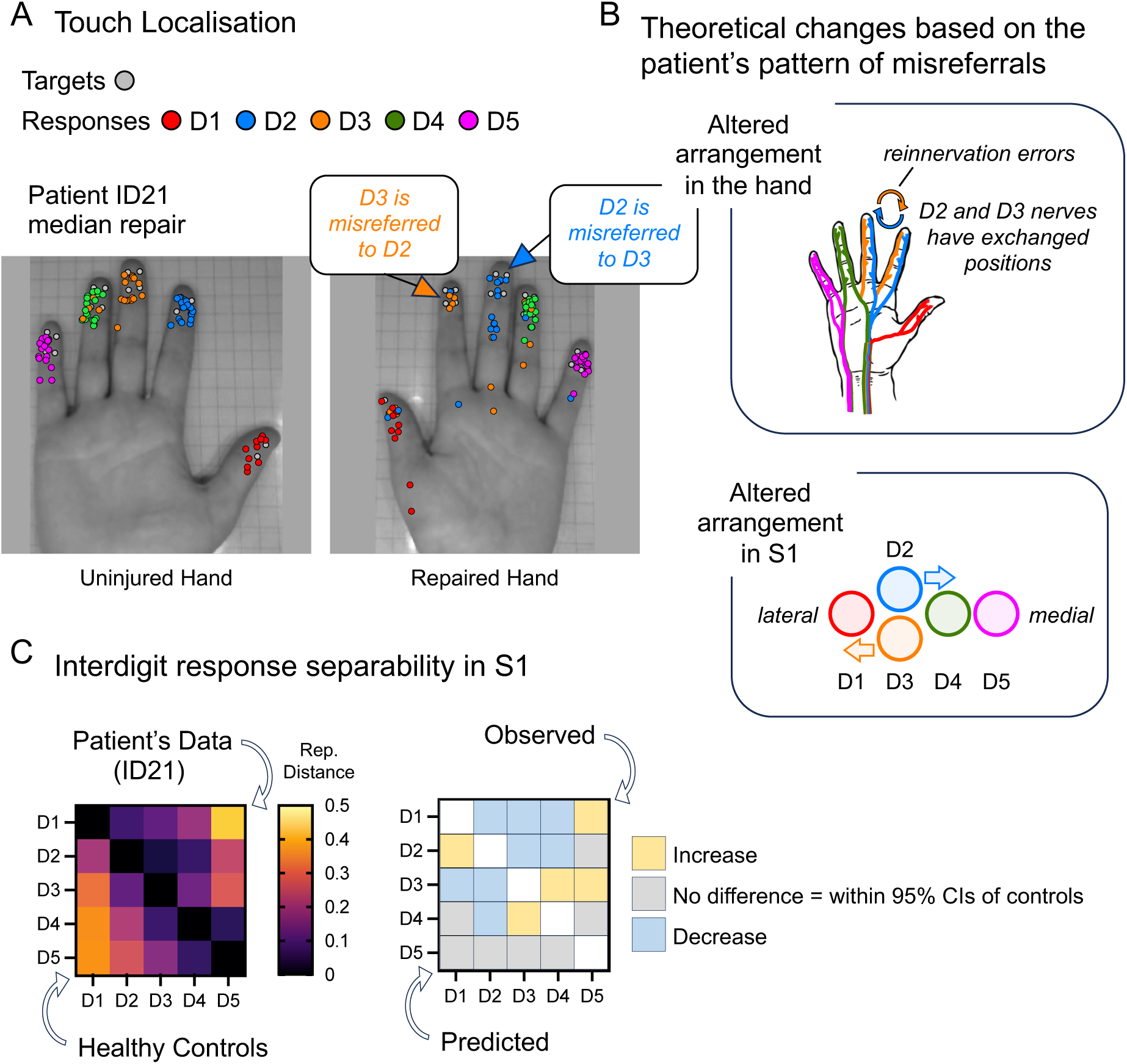
Frequent misreferrals between digits may reflect reinnervation errors. **A:** Locognosia results from patient ID21. Targets (grey circles) and responses (coloured circles) are overlaid on the photographs of the hand. The colours indicate which digits were touched (see inset key). Touch of D2 was frequently misreferred to D3, and sometimes vice versa. This figure has been modified from Weber et al.^35^, licensed under CC-BY. **B:** Theoretical changes in the arrangement of nerves in the hand based on the patient’s pattern of misreferrals. Reinnervation errors may have resulted in an exchange between D2 and D3 nerves. This would predictably alter the arrangement of digit maps in S1. For example, D2 should ‘move closer’ to D4. **C:** Representational distances are shown in the heatmap, with patient ID21’s data above the reference diagonal and the mean of healthy controls below (left). Predicted and observed differences between the patient’s data and controls (right). The observed differences are above the reference diagonal and predicted differences are below. For each interdigit comparison, ‘increased’/‘decreased’ observed results indicates that the patient’s representational distances are above/below the 95% confidence intervals of healthy controls, respectively.

Next, we test whether RDM typicality relates to sensory Rosen scores. The Rosen scale remains the most widely used clinical measure of functional impairments after median/ulnar nerve repairs^3,42^ (see Methods: *Sensory Rosen scores*), and all 21 patients completed testing. We find no reliable evidence for a relationship between sensory Rosen scores and RDM typicality (r_s_(19)=0.05, p=0.835; 95%-CI, [−0.40,0.48] BF_10_=0.35; Supplementary Fig. S9).

We then conduct exploratory correlations between RDM typicality and (i) the Sollerman subtests (within the sensory Rosen scale), (ii) patient-reported disability (DASH) and (iii) patient-reported pain (McGill)—both described in Methods: *Patient-reported outcomes*—and (iv) time since repair. We find no credible evidence for associations with RDM typicality across these analyses (Supplementary Fig. S10). Altered S1 maps do not reliably correlate with sensorimotor impairment (Sollerman), self-reported disability (DASH) or pain (McGill), or time since repair.

## Discussion

Seminal studies in non-human primates showed that forelimb nerve transection and repair dramatically alters the otherwise orderly and highly conserved organisation of the digit maps in primary somatosensory areas 3b and 1. These discoveries helped to establish the principle that the brain’s functional organisation depends on continued reciprocal interactions with the body, and laid the foundations for contemporary rehabilitation practice in nerve repair. Despite these profound influences, whether similar brain changes occur in humans remains uncertain.

Here, we directly test this question. Using high-resolution fMRI, we characterise the fine-grained organisation of digit responses in the primary somatosensory cortex (S1) in 21 patients with surgically repaired hand nerve transection injuries and 30 healthy controls. Cross-validated Mahalanobis distances show that overall separability of digit responses in S1 is preserved after nerve repair: mean representational distances do not differ between patients and controls. However, the internal structure of these distances is systematically altered. Compared to controls, patients exhibit a distorted representational geometry, indicating changes in the fine-grained organisation of digit maps. The pattern of interdigit separability, defined by the RDM structure, is altered in the hemisphere contralateral to the repaired hand (*S1-patients-repaired*). These alterations are accompanied by increased BOLD responses, despite definitive tactile impairments.

Consistent with altered digit map structure, measures of RDM typicality also reveal significant departures from the normative pattern. Surprisingly, typicality is also reduced in S1 of the uninjured hand (*S1-patients-uninjured*). Because bilateral changes are unexpected, we tested whether reduced typicality in patients simply reflects noisier data. Cross-validated within-subject reliability analyses provided no credible evidence for increased measurement noise in patients relative to controls. Decreased RDM typicality in S1 of the repaired hand also depends on which nerve(s) were repaired, showing significantly greater departures from normal after median or combined median–ulnar repairs than after isolated ulnar repairs. This specificity cannot be attributed to random noise.

Altogether, our findings support the conclusion that hand nerve transection and repair alters the fine-grained organisation of digit maps in S1. This is consistent with our second hypothesis (***H_2_ decreased typicality***) and supporting observations from animal models. Following forelimb nerve transection and repair in non-human primates, the typical orderly arrangement of S1 digit maps is transformed^9–12^. The normative pattern of skin-to-cortex adjacencies is altered, and many S1 neurons now also respond to touch at multiple discontinuous locations on the hand. These ‘multi-field’ neurons are found within the cortical zone of the repaired nerve and can have up to five different response fields on the hand, some of which may span multiple digits. Such changes provide a plausible neural basis for the distorted patterns of interdigit response separability that we find in *S1-patients-repaired*.

Peripheral reinnervation errors may explain these effects. Since nerve regeneration is unguided, sprouting fibres establish new connections ‘blindly’, changing the branching structure of nerves in the hand^6–8^. Digital nerves may reroute or exchange between digits. Unusual ramification of branching nerve fibres^5,6,43^ could also explain the emergence of ‘multi-field’ neurons. Afferent inputs normally conveying information from spatially continuous regions of skin may now communicate signals from spatially disparate regions^34^. These kinds of changes in peripheral input structure may explain the changes in digit-map structure observed in *S1-patients-repaired*.

It is useful to situate this account within the context of recent considerations regarding the limits of cortical reorganisation^44^. A peripheral rewiring interpretation implies that “altered maps” need not signal cortical reorganisation per se. The cortical circuitry itself may remain largely stable, with changes in map organisation reflecting a remapping of inputs rather than cortical plasticity. Nonetheless, the digit maps in S1 have changed.

This framing also helps reconcile our results with evidence for the stability of hand maps following deafferenting injuries such as amputation^45–47^ or spinal cord injury^48^. In both cases, the digit map organisation remains within the normative range, suggesting that cortical topography is robust to the loss of input. Nerve transection and repair, in contrast, restores input continuity via nerve regeneration. It is this inherently imperfect, unguided regeneration process that alters the hand-to-brain mapping, driving the distorted S1 digit-map structure we observe.

Of course, mechanisms of cortical plasticity may also be at play. We find elevated BOLD responses in *S1-patients-repaired*, a result consistently reported across previous investigations^24,25,27,28,30,31^. Such increases are unlikely to originate from the periphery: after nerve repair, tactile afferents may be reduced in number and sensitivity, and there is no evidence for enhanced peripheral drive^49^.

Instead, elevated BOLD effects are best explained by cortical disinhibition^25^. Animal studies reveal reduced inhibitory tone and homeostatic scaling of excitability, consistent with a high-plasticity state^50–54^. These processes may both preserve stability under altered inputs and permit adjustments, potentially enabling multiple perceptual/receptive fields to co-exist^55^. In nerve repair, such mechanisms may support adaptive refinements to the changes in organisation imposed by peripheral rewiring. Elevated BOLD responses in *S1-patients-repaired* may therefore mark an ongoing ‘attempt’ to stabilise altered circuitry, consistent with the principles of homeostatic plasticity^50^, while simultaneously enabling further change.

Our findings may also reflect use-dependent plasticity driven by compensatory changes in everyday hand use. In healthy controls, the representational geometry of digit maps in S1 reflects not only cortical topography but also the probabilistic patterns of finger co-use in daily actions^22^. Short-term manipulations in humans suggest that altered hand-use behaviour can reshape somatotopy^56^. Hand use is likely to change after nerve injury^4,57^, and this may alter digit map structure. This account may also explain why we observed reduced RDM typicality bilaterally. The pattern of hand use may have also changed for the uninjured hand.

We are unable to critically evaluate this interpretation. Real-world hand use was not directly measured; our clinical tools provide limited insight. Sollerman subtests involve goal-directed tasks but yield only coarse metrics (task completion time and subjective ratings of performance) and assess only the repaired hand. DASH captures self-perceived difficulties with activities of daily living, agnostic to how tasks are performed. Addressing this gap will require quantitative assessments of naturalistic hand use, ideally through ambulatory monitoring in real-world settings, with both hands tracked in parallel. Because changes in S1 of the uninjured hand were unexpected, these results should be interpreted cautiously.

We are surprised to find no credible evidence that altered S1 organisation is associated with impairments in touch localisation or broader functional outcomes (Sensory Rosen scores). Impaired locognosia in patients and reorganised S1 topography in animals exhibit common characteristics—restricted occurrence within nerve-injured zones and high individual variability^10,41^—and reinnervation errors may explain both phenomena^21,33,34^.

One limitation is that while our fMRI methods capture between-digit organisation, our locognosia protocol is optimised for the measurement of within-digit errors and less sensitive to between-digit errors (misreferrals). Adaptive psychophysical protocols reveal patterns of misreferrals in healthy controls that appear to reflect S1 organisation^58–61^. Future work may adopt these protocols, or complementary methods targeting interdigit perception^56^. Alternatively, ultra-high field fMRI may be used to map intradigit organisation^62,63^

Contemporary models of functional recovery in nerve repair place a strong emphasis on the brain and its capacity for change^4,13,64–70^. Our study validates a core assumption of these models—nerve repair alters the cortical maps of the hand—yet reveals no clear evidence that these changes bear directly on functional recovery. While this may reflect insensitive behavioural assays, or other study limitations, this also raises the possibility that such alterations may be less detrimental than often assumed. From this perspective, recovery may not depend on restoring canonical S1 organisation but refining and stabilising the altered mapping. Rehabilitation strategies such as sensory re-education therapy may therefore function less to “halt” or “reverse” cortical changes than to guide the system toward a stable and functional equilibrium.

Again, this framing resonates with the ideas on homeostatic plasticity^50^ and constrained cortical reorganisation^44^ discussed above. The distributed nature of digit representations in S1^71–73^ may provide a scaffold for adapting to altered inputs, helping to reduce the functional costs of distorted topography. Nonetheless, changes to the normally continuous skin-to-cortex adjacencies—the spatially ordered topography of the hand—are still likely to have functional consequences^13,74–76^. Our data provide no conclusive insights on this front. Clarifying the functional impact of altered S1 organisation remains a critical challenge, with implications not only for basic science and rehabilitation practice, but also for emerging neural prosthetic and brain–machine interface technologies that must restore body–brain communication through altered input mappings^77,78^.

In summary, we show that hand-nerve repair significantly alters the functional organisation of S1, yet the clinical relevance of these changes remains unclear. Given the inherent heterogeneity of peripheral nerve injuries and the complexity of factors influencing patient recovery^67^, future research will benefit from longitudinal measures and advanced modelling approaches to determine whether—and how—brain changes relate to patient outcomes. Incorporating direct measures of peripheral rewiring (e.g., using microneurography^34,79^) and quantitative assessments of real-world hand use may help disentangle bottom-up from use-dependent contributions to altered cortical organisation. Answering these questions will be crucial for linking mechanistic insights to meaningful clinical practice.

## Methods

### Participants

Procedures were approved by the Bangor University local Ethics Board and by the NHS ethics, Wales Research Ethics Committee 5, (REC reference: 16/WA/0157). All participants gave informed written consent according to the Declaration of Helsinki. Participants received financial compensation.

#### Patients

Twenty-one patients completed testing (see Supplementary Table S1 for demographics and standardised clinical assessment scores). Eighteen patients had complete transection injuries: ten ulnar, three median, and five ulnar and median (‘both’). Three patients had incomplete transection injuries of the median nerve (labelled ‘part median’ in Fig. 7). One patient’s injury was due to self-harm. This patient was deemed mentally stable when tested. All other injuries were of traumatic origin. All patients were injured in adulthood.

Surgical repair was performed to restore continuity across the injury site. Following this, axonal regeneration proceeds spontaneously and, as far as is currently understood, without topographical guidance—an inherently random process that can lead to reinnervation errors (see Fig. 1B). Precise timing of surgical repair relative to injury was not available. Most patients underwent repair within 4 days of injury.

Ten of the 21 patients sustained injuries to their dominant hand. We did not explicitly assess changes in hand dominance following injury, and no participants spontaneously reported switching handedness.

#### Healthy controls

Thirty participants completed MRI testing (age range: 19—63 years; mean age: 31.9 years; 13 female, 17 male). Two participants were left-handed.

For locognosia testing, the control group was partly different. In total, 33 participants completed locognosia (age range: 19—63 years; mean age: 30 years; 13 female, 20 male): 28 of these also completed MRI, while 5 were additional participants who did not undergo MRI.

### MRI

A Philips 3T Ingenia Elition X MRI scanner with a 32-channel headcoil was used. Functional scans involved vibrotactile stimulation of the distal digit pads of the hand using custom-built stimulators (Supplementary Fig. S1). Each digit was stimulated seven times per scan, with each hand tested separately. Stimulation lasted 4s with bursts at 30Hz (500ms on, 500ms off). Scans began and ended with rest periods, involving fixation only, and included five additional rest periods (13-16s) interspersed throughout. Condition order was pseudo-randomised within scans, controlling for condition history, and sequence order was counterbalanced between participants. Total run duration was four minutes.

Stimulators were held using a custom-built base, which followed the hand’s curvature and allowed adjustment to accommodate different hand sizes. The actuating terminus of each stimulator was aligned with the whirl of each digit. In cases where patients had difficulties straightening their fingers, tape was used to fixate the stimulators. Participants fixated a cross on a screen viewed through mirrors and were instructed to attend to each stimulation.

Stimulator amplitudes were standardised for all participants. Mechanical measurements were taken prior to the study to set the displacement amplitude of ‘stronger’ stimulators to approximate that of the ‘weakest’ stimulator.

Before each scanning session, participants were given time to familiarise themselves with the stimulators. This included feeling a structured sequence of stimulation events to each digit, delivered in thumb-to-little-finger order. Participants were asked to report the order in which they perceived stimulation, confirming that stimulation of a given digit produced a correctly localised percept. This procedure was repeated inside the scanner, after participants were comfortably positioned. These checks ensured that stimulators were correctly aligned, and confirmed that each finger was being stimulated as intended.

During this process, patients were also encouraged to describe the perceptual quality of stimulation delivered to their repaired digits, including any deviation from normal touch. All patients reported feeling stimulation at all five digits, and none reported pain. Some patients spontaneously noted that although each finger was correctly stimulated and perceived, the sensation was not always localised to the fingertip, but rather was perceived as more proximal or diffuse. No patient reported feeling stimulation on the wrong digit; all were able to report the correct stimulation sequence. Some patients also spontaneously described touch on their reinnervated skin as “unusual”—a generalised phenomenon not specific to the vibrotactile stimulation used during scanning (see pain/discomfort subtest of the Rosen score; Supplementary Table S1).

Following each functional run, participants were asked to confirm that stimulation had been felt at each digit throughout the entire run. These checks were used to detect any stimulator displacement or malfunction. Together, these procedures minimise the likelihood of improper stimulation delivery and ensure that the digit-specific stimulation was reliably perceived throughout.

Scanning was completed in two sessions, one per hand, each comprising five functional scans and a field map scan. The first session included an anatomical scan. Each session took approximately 30 min.

The repaired hand was always tested first to prioritise its measurement in case of early discontinuation, which occurred with one patient due to anxiety. This patient completed four scans with their repaired hand and three scans with their uninjured hand. All other participants completed all scans. For healthy controls, hand order was counterbalanced.

#### Scan parameters

Functional scans were taken at an in-plane resolution of 2 x 2 mm (T_2_*-weighted echo-planar sequence, TR = 2000 ms, TE = 30 ms, flip angle = 90°, 58 contiguous interleaved slices with 2 mm thickness, matrix size = 112 x 110, multiband = 2, bandwidth = 1905 Hz/pixel). The T_1_-weighted structural scan was acquired using an MP-RAGE pulse (TR = 18 ms, TE = 3.43 ms, flip angle = 8°, voxel size = 1 mm isotropic, field of view = 225 x 225 x 175 mm). Field maps were acquired using a double gradient echo sequence. Field maps were not collected for the first six participants due to technical problems. These participants were healthy controls.

#### MRI analysis

MRI data were analysed using FSL (v6.0, http://www.fmrib.ox.ac.uk/fsl/)^80^. Non-brain structures were removed using BET. Head movement was processed using MCFLIRT. EPI unwarping was performed using FSL PRELUDE and FUGUE. High-pass filtering (90s cut-off) was applied. Grand-mean intensity normalisation of the entire 4D dataset was applied by a single multiplicative scaling factor (FEAT default).

Functional data were registered to structural scans using *epi_reg*, which performs an initial 6 degree-of-freedom rigid-body pre-alignment using the *corratio* cost function, followed by refinement with boundary-based registration^81^, also using 6 degrees of freedom. Field map-based unwarping was applied during this process to correct for susceptibility-induced EPI distortions, using a B0 field map and magnitude images (echo spacing: 0.0003677 seconds; phase encoding direction: y; EPI echo time: 30 ms; signal loss threshold: 10%).

The hemodynamic response function was modelled as independent predictors locked to the onsets and set to the durations of each vibrotactile stimulation event per condition: D1, D2, D3, D4 and D5. Additional covariates included single-point predictors for high-motion outliers identified using fsl_motion_outliers, using default settings. This procedure computes the root mean square (RMS) intensity difference between each volume and a reference volume, and flags volumes as outliers if their RMS value exceeds the box-plot cutoff, defined as the 75th percentile plus 1.5 × the interquartile range.

First-level contrasts of parameter estimates were calculated for D1 > rest, D2 > rest, D3 > rest, D4 > rest, and D5 > rest. Second-level analyses were performed for each participant by combining first-level analyses (i.e., all functional runs per hand) using a fixed-effects model.

#### Regions of interest

S1 regions of interest (ROIs) were defined using the functional probabilistic atlas developed by O’Neill et al. (2020)^35^. The atlas is based on independent data from 22 healthy participants acquired at 7T MRI (see^35^ for complete methods). Briefly, vibrotactile stimulation was delivered to each fingertip using a travelling wave paradigm, comprising 8-12 stimulation cycles per functional scan, with two scans per hand (alternating forward and backward sequences). For each participant, digit-specific activation maps were coherence thresholded at p < 0.05 (uncorrected), restricted to subject-specific FreeSurfer anatomical labels of Brodmann areas 3a, 3b, 1, and 2, and combined across subjects to form full probabilistic maps. Five maps were generated per hemisphere, corresponding to each digit (D1-D5). The value at each vertex represents the proportion of subjects exhibiting significant thresholded activation for that digit. Note that the maximum probability differs across digit maps due to inter-subject functional-anatomical variability.

To define ROIs for the current study, the full probabilistic digit maps of the O’Neill atlas were first projected from FreeSurfer fsaverage surface space onto each participant’s individual cortical surfaces, and then to their native volume space. For each hemisphere, a whole-hand ROI was created by thresholding each of the five full probabilistic digit maps at a minimum probability value of 0.046 (just greater than the equivalent of one participant), summing these thresholded maps, and binarizing the combined result. This liberal threshold was chosen to increase the likelihood of accurately capturing the location of S1 across our participants, given that the atlas is derived from an independent dataset and there is considerable interparticipant variability in functional anatomy. The resulting binarized whole-hand mask comprises our S1 ROIs per participant per hemisphere.

Supplementary Figure S2 shows the S1 ROIs for two participants.

Our original analyses defined S1 ROIs manually. For each participant, the central sulcus was traced 10 mm above and below the hand knob, aiming to restrict selection to the sulcus itself and avoid pre- and postcentral gyri. We subsequently adopted the atlas-based approach following reviewer feedback. This provides a more principled and reproducible basis for ROI definition while still accommodating interindividual anatomical variability. For transparency, data tables from the original manual ROI analyses are available in our public repository; statistical results were largely consistent across ROI methods.

### Measuring digit map separability

The separability of interdigit responses was measured using multivariate representational distances^82^. Representational distances reflect the level of dissimilarity between pairs of unthresholded digit response patterns. Greater values indicate greater separability.

In our original analyses, we included Dice overlap coefficients as a complementary univariate measure of digit-response separability. Dice coefficients quantify the spatial overlap of positive-thresholded digit activation maps, with higher values indicating greater overlap^46,83^. We subsequently removed Dice analyses from the main manuscript for several reasons.

First, multivariate representational distances provide a more robust and better-validated approach: they incorporate noise estimates, have a principled zero baseline, and yield greater reliability^23^. Consistent with this, Dice results show considerably higher interparticipant variability.

Second, Dice has inherent limitations that complicate interpretation. When activation is weak, Dice values approach zero—not necessarily because of true spatial non-overlap, but because of insufficient signal. In the extreme, if one contrast yields no suprathreshold voxels, Dice is zero even if underlying distributions overlap. Conversely, when activation is widespread, Dice can approach 1.0 even if response peaks differ.

These issues are compounded when comparing groups that differ in response robustness—whether measured by signal amplitude (%-BSC) or activation volume (thresholded voxel counts; see Methods: *Activation Volume*). In such cases, Dice values conflate overlap with responsivity. Indeed, Dice coefficients strongly correlate with %-BSC (Supplementary Fig. S11A) and activation volume (Supplementary Fig. S11B). This indicates that Dice values are strongly driven by response robustness, making it difficult to determine whether group-level differences in Dice reflect true differences in map overlap or differences in signal strength. By contrast, representational distances and RDM typicality show no such correlations (Supplementary Fig. S6)

Nonetheless, in our revised analyses—using surface-based atlas ROIs—Dice and representational distances yield statically equivalent results. For completeness and transparency, Dice results are reported in Supplementary Table S2, accompanied by interpretational caveats.

#### Representational distances

Representational distances were calculated for each pair of digit response maps as cross-validated squared Mahalanobis distances (using the toolbox from Nili *et al*^36^ adapted for FSL^84^). For each digit response map, beta values from all voxels within the contralateral S1 ROI formed the complete pattern of responses. This includes all (unthresholded) beta values; the full-spectrum of positive- and negative-going fMRI responses. Extracted betas were then prewhitened using the voxel-wise residuals, and each run was used as an independent cross-validation fold to compute average (cross-validated) Mahalanobis distances across folds^82^. If two response patterns differ only in noise, their representational distance will be zero. Computing all pairwise interdigit distances generates a representational distance matrix (RDM) with 10 unique measures.

### Evaluating hypotheses

Representational distances were computed for each participant and hemisphere ROI. Predictions regarding functional changes due to nerve repair are specific to S1 contralateral to the repaired hand, *S1-patients-repaired*. To evaluate our hypotheses, we performed the following comparisons: (1) we compared *S1-patients-repaired* against measures from healthy controls, *S1-controls*; (2), we compared *S1 patients-repaired* against measures taken from the patient’s uninjured hand, *S1-patients-uninjured*; (3) we tested *S1-patients-uninjured* against *S1-controls* where no differences were predicted.

Data from healthy controls were extracted from the contralateral S1 ROI of each hemisphere and averaged. Control data showed no significant differences between dominant and non-dominant hands (Supplementary Table S3).

#### Statistical analysis

All tests were two-sided with α = 0.05. Tests for normality were performed using the Shapiro–Wilk test. When normality was violated (p < 0.05), we used the non-parametric counterpart to the planned test. Between-group contrasts used independent-samples t-tests or Mann–Whitney U tests; within-participant contrasts used paired t-tests or Wilcoxon (W) signed-rank tests. Associations were examined with Pearson’s r when normality held and Spearman’s rank correlation (denoted as r_s_) otherwise.

All families of related comparisons were corrected for multiple comparisons using the Holm–Bonferroni procedure, with α set at 0.05. Corrected p-values are reported in the main text when uncorrected p-values were ≤ 0.3. In all other cases, uncorrected p-values are reported.

Non-significant results were further examined using Bayesian hypothesis testing. Bayes factors (BF_10_) were calculated using JASP^85^. We report Bayes factors (BF_10_), calculated using default JASP settings (Cauchy prior width r = 0.707), to quantify evidence relative to the alternative hypothesis. A BF_10_ < 1/3 is interpreted as moderate evidence in favour of the null.

Factorial ANOVAs were used to evaluate: (1) RDM structure: Group/Hemisphere (2 levels) x digit-pair (10 levels) (see: Evaluating H2: RDM structure is altered after nerve repair; Fig. 4); (2) RDM typicality, Hemisphere (2 levels) x patient-subgroup (2 levels) (see: Decreased typicality on the repaired side depends on the nerve(s) repaired; Fig. 5C). Assumption checks for ANOVAs included visual inspection of Q-Q and residual-versus-fitted plots to assess normality and residual structure, and inspection of homoscedasticity plots to evaluate variance equality across groups. All post-hoc comparisons of significant interactions were conducted as above, using either parametric tests (paired/independent sample t-tests) or non-parametric (Wilcoxon/Mann-Whitney) equivalents, Holm–Bonferroni corrected for multiple comparisons.

For non-significant results, we report inclusion Bayes factors (BF_incl_) computed using JASP’s default Bayesian ANOVA settings (uniform model prior; Cauchy priors on fixed effects r = 0.5, random effects r = 1; marginality enforced).

### H_1_ decreased separability

Nerve repair should decrease the separability of interdigit responses in S1. Smaller representational distances are predicted.

#### Repair-zone-specific analyses

Previous findings from monkey neurophysiology indicate that cortical changes in areas 3b/1 are specific to digit response maps within the zone of the repaired nerve(s)^9–12^. We therefore repeated tests of H_1_ using only those representational distances computed based on comparisons between maps within the nerve repair zone.

For patients with isolated median nerve repairs, this included D1–D2, D1–D3, D1–D4, D2–D3, D2–D4, and D3–D4 comparisons. For isolated ulnar nerve repairs, this included D4–D5. For patients with combined median and ulnar nerve injuries, all pairwise digit comparisons comprised the repair zone and were included (see Fig. 3C-ii for illustration). For *S1-patients-uninjured*, mean representational distances were computed from the corresponding homotopic digit pairs.

To ensure fair patient–control comparisons, we organised the control data to match the distribution of nerve injury types in the patient cohort. Without this step, group-level estimates would be based on different sets of digit-pair comparisons. Specifically, six patients had isolated median nerve injuries (∼29%), so nine controls were assigned to a “median-nerve-repair” subgroup (∼30%) and their data restricted to D1–D4 comparisons. Ten patients had isolated ulnar nerve injuries (∼48%), so 14 controls were assigned accordingly (∼47%) and their data restricted to D4–D5. The remaining seven controls were matched to patients with combined median-and-ulnar injuries, and their data included all pairwise digit comparisons. Control assignment within each subgroup was otherwise random.

### H_2_ decreased typicality

Nerve repair is expected to transform the typical arrangement of digit responses maps in S1. Patients should deviate from the stereotypical arrangement of interdigit separability observed in healthy controls.

#### RDM structure

To directly test whether nerve repair alters digit-map structure, we conducted a mixed ANOVA with the factors group (*S1-patients-repaired* vs. *S1-controls*) and digit-pair (10 levels; all possible digit-pair comparisons comprising the RDM structure). A significant group x digit-pair interaction would indicate systematic alterations in interdigit representational geometry following nerve repair, directly supporting ***H_2_ decreased typicality***.

We additionally conducted two complementary tests: a repeated-measures ANOVA comparing the two hemispheres of patients (*S1-patients-repaired* vs. *S1-patients-uninjured*), and a second mixed ANOVA comparing the uninjured hemisphere of patients to healthy controls (*S1-patients-uninjured* vs. *S1-controls*).

*Visualisation of RDM structure*. For illustrative purposes (Fig. 4), we applied within-subject normalisation and multidimensional scaling (MDS).

▪ Normalisation (Fig. 4A-E): For *S1-controls*, *S1-patients-repaired*, and *S1-patients-uninjured*, representational distances were normalised by dividing each distance by the participant’s mean distance per condition. This highlights the relative structure of the RDM while removing across-subject differences in absolute distance scale.
▪ Multidimensional scaling (MDS) (Fig. 4F): To visualise representational geometry per condition, mean RDMs were projected into two dimensions using classical MDS^86,87^. This procedure preserves relative dissimilarities between digit representations Formally, the squared dissimilarity matrix was double-centred, decomposed, and reduced to the two leading eigenvectors, which defined the 2D coordinates.

#### RDM typicality

Typicality scores were computed for each participant by comparing their interdigit separability patterns—RDMs—against the group mean pattern in healthy controls, generating an r-value for each measure (using Spearman’s rho; r_s_). The r_s_-values reflect the degree of correspondence between an individual’s pattern and the control group mean. High typicality indicates close agreement with the normative pattern, while low typicality indicates divergence.

To estimate typicality in controls, we used a leave-one-subject-out analysis, comparing each individual’s RDM against the mean RDM of all other controls.

#### RDM reliability

To evaluate whether reduced RDM typicality in patients could reflect noisier estimates of representational geometry, we assessed within-subject reliability across imaging runs. For each participant and hemisphere (i.e., imaging session), we computed the Pearson correlation coefficient between the RDM from each held-out run (test set) and the mean RDM from the remaining runs (train set). Correlations were then averaged across folds to yield a single reliability score per subject per hemisphere. This conservative cross-validation procedure provides an estimate of internal consistency that is independent of run-specific noise.

### Percent BOLD signal change

For each participant, mean contrast of parameter estimate (COPE) values were extracted from S1 ROIs using FSL *featquery*. COPEs were averaged across runs (higher-level *gfeat* analysis) and converted to percent BOLD signal change (%-BSC) values. Values were calculated separately for contralateral and ipsilateral stimulation relative to each ROI.

Whole-hand analysis: COPEs were averaged across the entire S1 hand ROI in volume space, using the same atlas thresholding procedure applied in all ROI analyses (see Regions of interest).

Repair-zone-specific analysis: ROIs were defined according to the repaired nerve(s), using a more selective (half-maximum) threshold applied separately to each digit map in the probabilistic atlas. Because each digit map of the atlas has a unique maximum (reflecting peak overlap across participants), the half-maximum threshold was applied relative to that map’s own maximum. Injury-specific ROIs were defined as follows: median repairs (D1–D4), ulnar repairs (D4–D5), and combined median and ulnar repairs (all five digits). %-BSC values were then extracted separately for contralateral and ipsilateral stimulation. Within each patient subgroup, the signal was averaged across the relevant injured digits (e.g., for ulnar repairs, values from D4 and D5 were averaged within the D4–D5 ROI).

### Activation volume

Activation volume was computed as an additional descriptive measure of response robustness. These analyses were motivated by reviewer concerns about whether each participant showed significant digit responses and how such variability might influence Dice results. Because activation volume is highly correlated with %-BSC, and %-BSC provides a more principled measure of response robustness, continuous and not dependent on thresholding, activation volume was not included in our main analyses. Results are reported in Supplementary Fig. S12 for completeness.

For each participant, digit-specific contrasts (D1–D5 > rest) were thresholded at z > 2.0 (uncorrected), restricted to the contralateral S1 ROI, and the number of suprathreshold voxels was computed. Volumes are reported both as raw voxel counts (1mm³ resolution; Supplementary Fig. S12A) and as proportions of total ROI volume (Supplementary Fig. S12B). While most participants showed robust activation for all five digits, low responsivity was observed for some contrasts in both patients and controls.

Group comparisons broadly mirror %-BSC results: activation volume is elevated for *S1-patients-repaired* relative to *S1-patients-uninjured*, but—unlike %-BSC—does not differ significantly from controls (Fig. S12C; see figure for full statistics). Importantly, activation volume in *S1-patients-repaired* does not correlate with representational distances or RDM typicality (Supplementary Fig. S6), confirming that our multivariate results are not confounded by variability in signal strength or activation extent. Finally, as with %-BSC, activation volume is strongly correlated with mean Dice coefficients (Supplementary Fig. S11C), further demonstrating that Dice values are confounded by response robustness, complicating their interpretation as measures of map overlap.

### Behavioural tests

#### Locognosia

Locognosia was measured using the Digital Photograph Method, detailed previously^41^. Nineteen of 21 patients completed testing (Supplementary Table S1).

Briefly, the participant’s hand was blocked from their view and points were marked on the volar surface using an ultraviolet (UV) pen (Supplementary Fig. S6). These points served as targets for touch stimulation. Two photographs were taken, one with and one without UV lighting, rendering targets seen and unseen, respectively. The experimenter used the photograph with visible targets to register their locations, and participants reported the felt position of each stimulation on the photograph with unseen targets.

We report two measures: (1) absolute error, the Euclidian distance between target-response pairs in millimetres; and (2) Misreferrals, responses made to an incorrect digit or to the palm of the hand. Misreferrals are excluded from the calculation of absolute error.

#### Sensory Rosen scores

The Rosen test is a standardised clinical assessment of hand function after median/ulnar nerve repair^3,42^. It comprises three independent domains—sensory, motor, and pain—with each domain scored separately. All 21 patients completed the sensory and pain domains; the motor domain was not tested.

Procedures followed standard protocols^88^. Touch detection thresholds used Semmes-Weinstein monofilaments; two-point discrimination used The Two-Point Discriminator (Exacta Precision & Performance, 2019 North Coast Medical, Inc.). Shape-texture-identification and the Sollerman test materials were produced in-house.

Importantly, while labelled the sensory domain, the sensory Rosen score is a composite measure that includes both low-level tests of tactile sensitivity—mechanical touch detection and two-point discrimination—and high-level tests of haptic processing (shape-texture-identification^89^) and manual function requiring fine movement dexterity (Sollerman^90^ subtests 4, 8, and 10). Thus, the sensory Rosen score reflects both sensory and motor aspects of hand function. Supplementary Table S4 provides patient sensory Rosen scores per subtest.

#### Patient-reported outcomes

Patients completed two self-reported questionnaires: the 30-item Disabilities of the Arm, Shoulder and Hand (DASH)^91^ and the Short-Form McGill Pain Questionnaire (SF-MPQ)^92^. The DASH yields a single disability score from 0–100, with higher values indicating greater disability. The SF-MPQ provides sensory and affective sub-scores, as well as a descriptor total (0–45); we report the descriptor total. Both questionnaires were administered in English and scored according to standard manuals.

#### Vibrotactile detection thresholds

We administered a vibrotactile detection–threshold test in a subset of participants (16 patients; 24 controls) using the same stimulators as in the MRI experiment. This assessment was originally included as a quality-assurance check to verify that the fixed stimulation intensity used during scanning was suprathreshold for each digit. This was important because absent or weak BOLD responses could, in principle, reflect either altered cortical responsivity or simply subthreshold peripheral stimulation.

Participants were seated comfortably with the tested hand resting palm-down. Digits were assessed one at a time using a single stimulator. Starting from zero, stimulus intensity was increased in fixed steps; at each level two pulses were delivered to minimise missed detections inflating the estimate. Participants indicated detection with a button press using the opposite hand. After a detection, intensity returned to zero before resuming the sequence. For each digit, the detection level was measured three times and averaged to yield the threshold.

Testing was performed after the MRI session. All patients detected stimulation at all digits of the repaired hand, and all thresholds were below the intensity used in the scanner, confirming that the fMRI stimulation was perceptible (Supplementary Fig. S5A). Because this was not part of the core protocol and testing days were lengthy, the procedure was de-prioritised once early data confirmed suprathreshold perception in patients; for this reason, thresholds were not collected in 5 of the 21 patients.

## Data availability

Summary data are available at github.com/weberetal/fMRI_PNR

## Acknowledgements

This work was supported by the Wellcome Trust (215186/Z/19/Z, to K.F.V.). We thank David McKiernan and James Naunton Morgan for technical support, including the design and construction of experimental materials. We are grateful to Rebecca Henderson for coordinating participant visits. We also thank Katja Kornysheva for advice on revisions and on the RDM reliability analyses, and Simon Watt for contributions to early discussions around the initial grant plans. We are especially grateful to the participants for their time and commitment, many of whom travelled long distances to take part.

## Author contributions

K.F.V., A.M. and V.L. conceived and designed the study. M.W. and R.Ti. collected and processed data. O.O., L.B., E.J., V.L. contributed clinical materials. V.L. and E.J. provided clinical oversight. M.W., R.Ti., M.A. and K.F.V. analysed data. M.W., A.M., F.M., S.F., M.A., E.J., V.L. and K.F.V. interpreted results. M.W., R.Ti., M.A. and K.F.V. prepared figures. M.W. and K.F.V. drafted the manuscript. M.W., A.M., F.M., R.Tu., S.F., M.A., V.L. and K.F.V. edited and revised the manuscript.

---

**Supplementary Figure S1.**
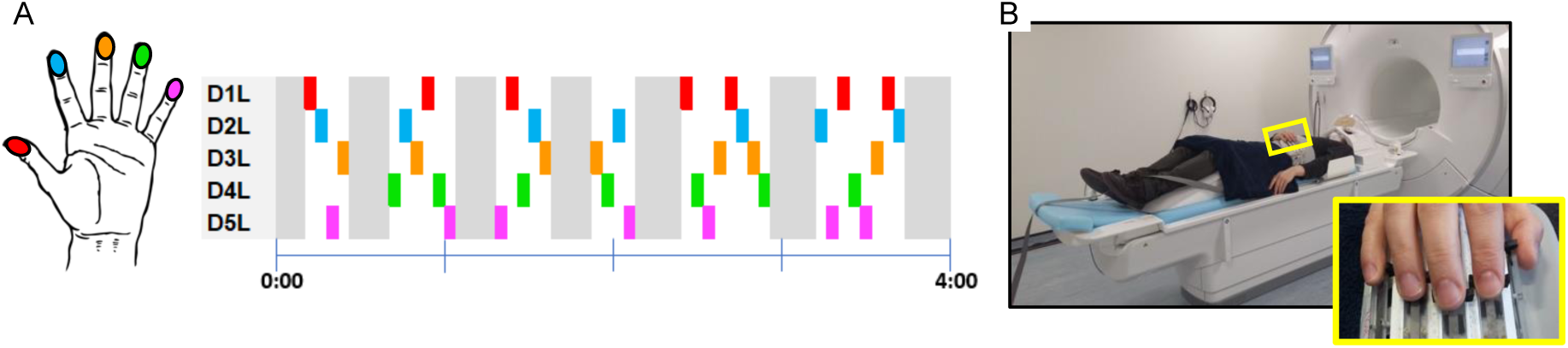
Functional MRI design and setup. **A:** An example of a single run. Each digit is stimulated seven times in the run, with periods of rest interspersed throughout. Each hand was tested separately. D1L = left thumb; D2L = left index finger; D3L = left middle finger; D4L = left ring finger; D5L = left little finger. **B:** An image of the participant setup in the scanner. The inset shows the piezoelectric stimulators.

**Supplementary Figure S2.**
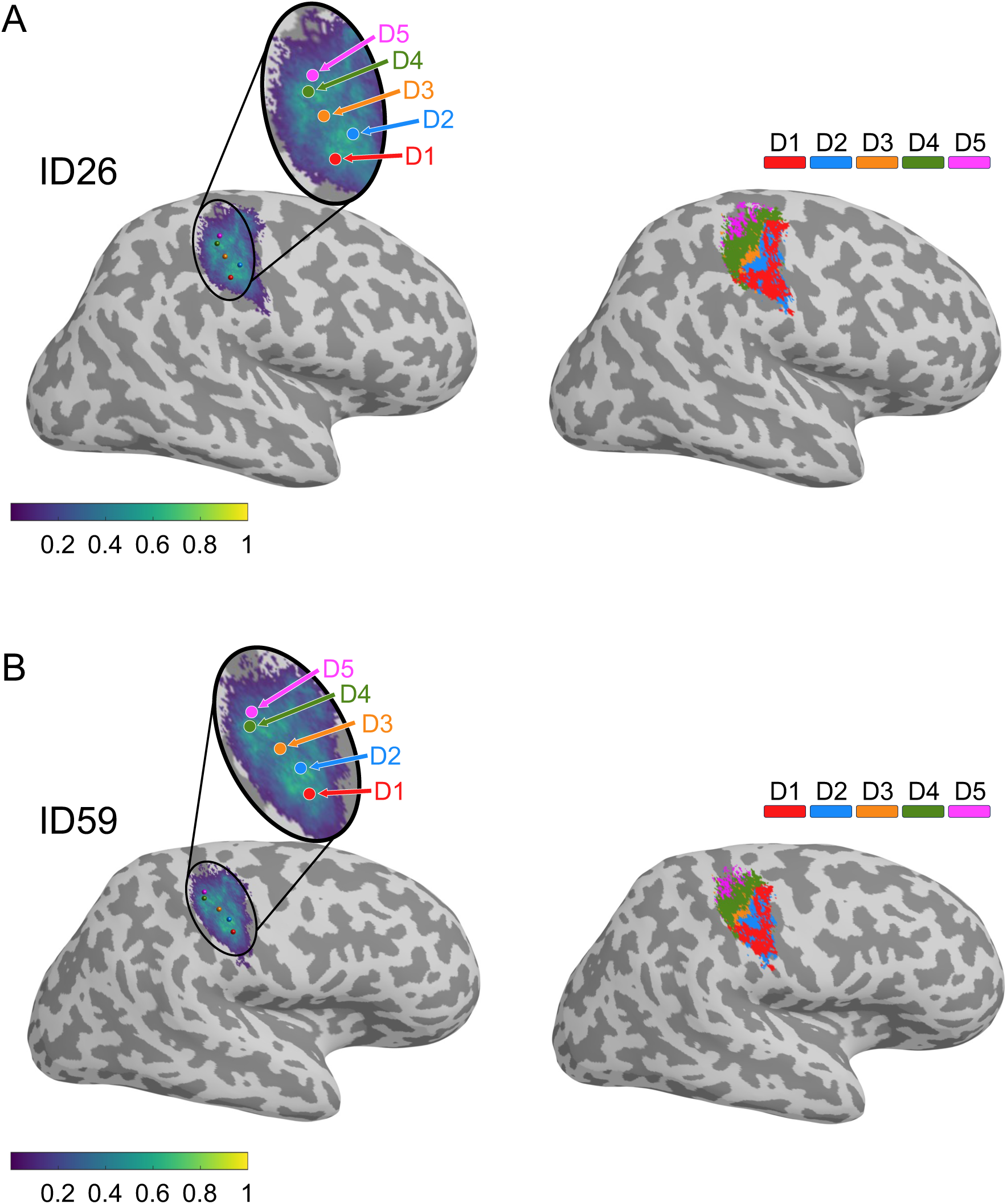
Atlas-based ROIs. Example S1 regions of interest (ROIs) derived from the probabilistic atlas of O’Neill, et al. (2020)^1^ for two participants (A: ID26; B: ID59). **Left**: the five digit (D1–D5) probabilistic maps overlaid, with peak participant-overlap locations per map shown as coloured spheres. The projected atlas corresponds to the posterior bank of the central sulcus and crown of the postcentral gyrus in each participant. **Right:** the same five digit-maps thresholded at a minimum probability of 0.046 (i.e., more than 1 of 22 participants) and binarized to create a whole-hand ROI. Colours indicate digit-specific ROIs after binarization.

**Supplementary Figure S3.**
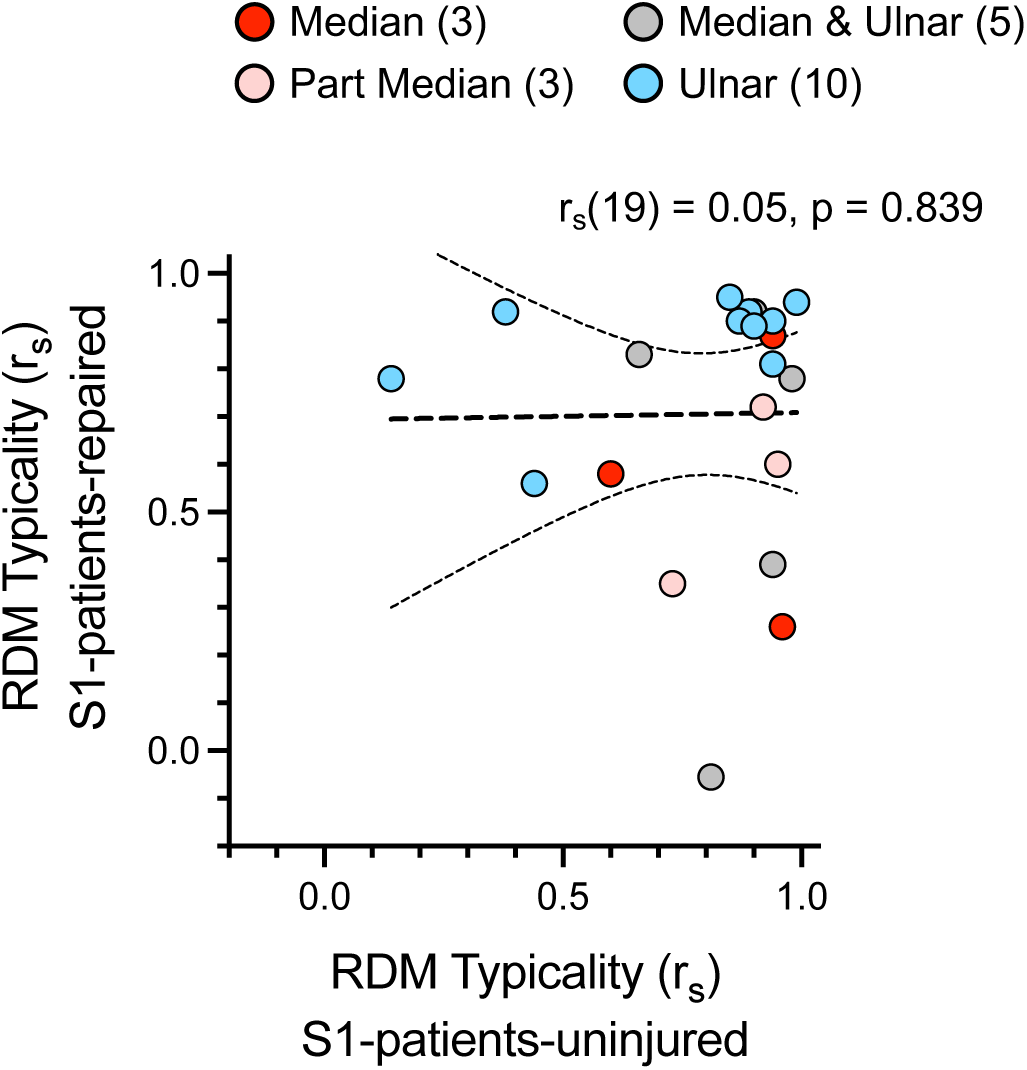
RDM typicality in patients does not correlate between hemispheres. Scatterplots showing RDM typicality for *S1-patients-repaired* versus RDM typicality for *S1-patients-uninjured*. No evidence for a relationship is found (r_s_(19)=0.05, p=0.839; 95%-CI, [−0.40,0.48]; BF_10_=0.30). These results are inconsistent with a mirrored interhemispheric remapping account in which altered S1 maps in the uninjured hemisphere reflect those of the repaired hemisphere. ‘Part median’ indicates patients with incomplete median nerve transection injuries.

**Supplementary Figure S4.**
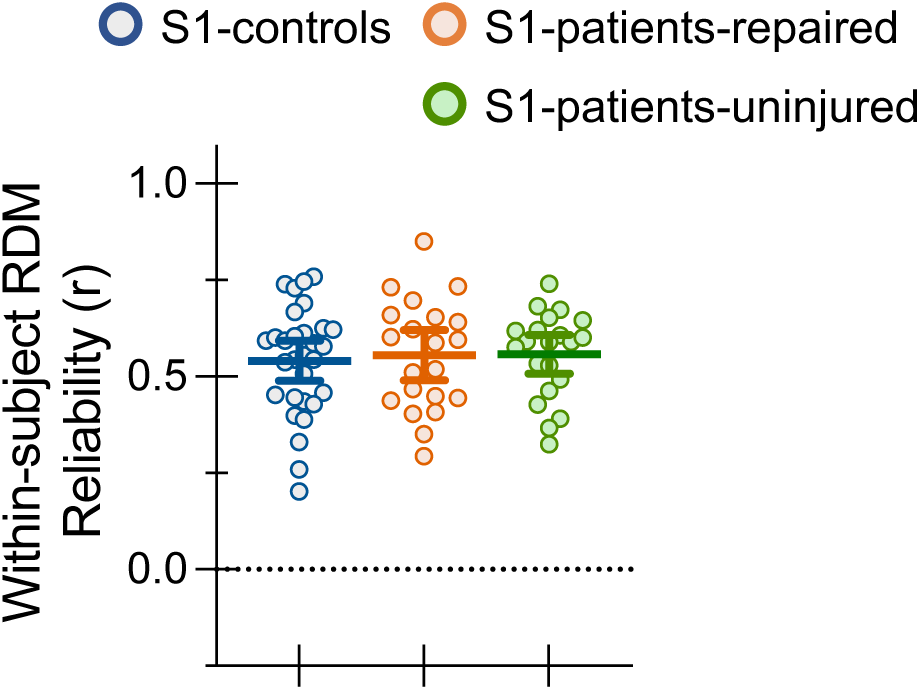
Within-subject RDM reliability. Reliability of RDM measures across imaging runs, for *S1-patients-repaired*, *S1-patients-uninjured*, and *S1-controls*. Reliability was computed by correlating the RDM from a held-out run with the average RDM from the remaining runs, repeated across all folds and averaged per participant. This provides a conservative test of internal consistency, since each comparison uses independent data folds. Reliability scores are modest overall, as expected for single-run analyses, but critically, they do not differ between patients and controls. Statistical results: *S1-patients-repaired* vs *S1-controls* (*t*(49)=0.35, p=0.727; η^2^=0.003; 95%-CI, [−0.07,0.09]; BF_10_=0.30); *S1-patients-uninjured* vs *S1-controls* (*t*(49)=0.46, *p*=0.648; η^2^=0.004; 95%-CI, [−0.06,0.09]; BF_10_=0.31); *S1-patients-repaired* vs *S1-patients-uninjured* (*t*(20)=0.07, *p*=0.945; η^2^ =0.0002; 95%-CI, [−0.08,0.08]; BF_10_=0.23). These results argue against the possibility that reduced RDM typicality in patients reflects increased measurement noise or instability across runs. Error bars are 95% confidence intervals.

**Supplementary Figure S5.**
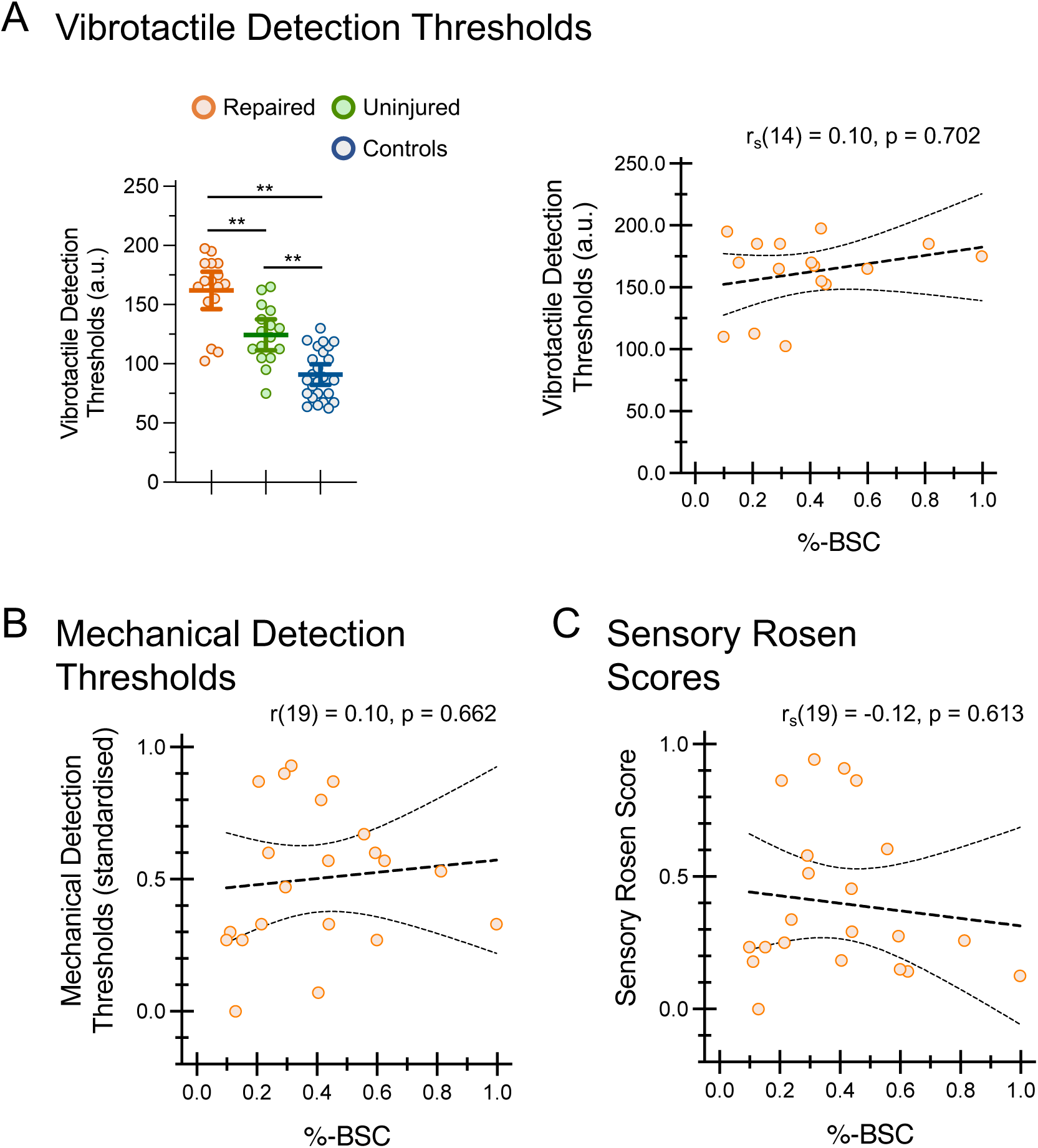
Percent BOLD signal change does not correlate with tactile sensitivity or sensory Rosen scores. Vibrotactile thresholds (see Methods: *Vibrotactile detection thresholds*). *Left*: group means for the repaired hand (N = 16), uninjured hand (N = 16), and controls (N = 24) (in arbitrary units of stimulator intensity). The repaired hand shows significantly elevated thresholds—i.e., reduced sensitivity—compared to both the uninjured hand and controls. Unexpectedly, the uninjured hand also shows reduced sensitivity relative to controls. Statistical results: repaired vs controls (t(39)=9.04, p<0.0001; *η*^2^=0.68; 95%-CI, [55.22,87.05]); repaired vs uninjured (t(15)=6.16, p<0.001; *η*^2^=0.72; 95%-CI, [24.52,50.48]); uninjured vs controls (unpaired t-test with Welch’s correction: t(28.48)=4.56, p<0.001; *η*^2^=0.42, 95%-CI, [18.53,48.73]). Asterisks indicate multiple-comparison-corrected significance for group comparisons (**p<0.001; two-sided). Error bars are 95% confidence intervals. *Right*: Scatterplots showing vibrotactile detection thresholds (repaired hand) versus %-BSC for *S1-patients-repaired*. No evidence for a relationship is found (r_s_(14)=0.10, p=0.702; 95%-CI, [−0.43,0.58]; BF_10_=0.40). **B-C:** %-BSC in *S1-patients-repaired* does not correlate with mechanical detection thresholds (r(19)=0.10, p=0.662; 95%-CI, [−0.35,0.51]; BF_10_=0.30) (B) or sensory Rosen Scores (r(19)=-0.12, p=0.613; 95%-CI, [−0.52,0.33]; BF_10_=0.31) (C).

**Supplementary Figure S6.**
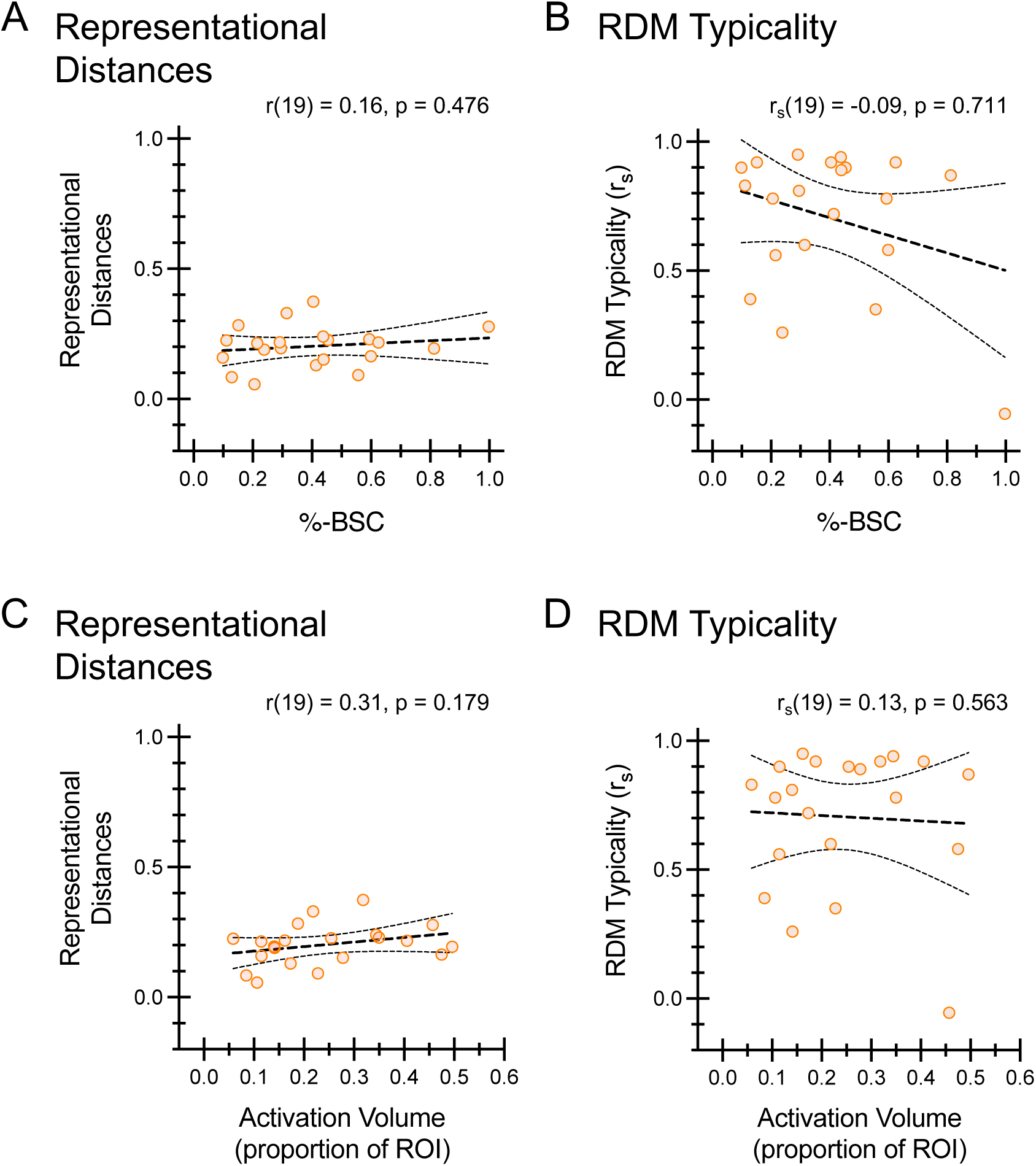
Percent BOLD signal change and activation volume do not confound RDM-based measures. **A-B:** Scatterplots showing the relationship between %-BSC and mean representational distances (A), and %-BSC and RDM typicality (B), in S1 of the repaired hand (*S1-patients-repaired*). Neither measure shows a significant correlation (A: r(19)=0.16, p=0.476; 95%-CI, [−0.28, 0.56]; BF_10_=0.34; B: r_s_(19)=-0.09, p=0.711; 95%-CI, [−0.51,0.37]; BF_10_=0.39). **C-D:** Same layout as A–B, showing the relationship between activation volume (number of suprathreshold voxels as a proportion of the total number of ROI voxels; see Methods: *Activation volume*) and mean representational distances (C), and activation volume and RDM typicality (D). Again, no significant correlations are observed. (C: r(19)=0.31, p=0.179; 95%-CI, [−0.16,0.65]; BF_10_=0.63; D: r_s_(19)=0.13, p=0.563; 95%-CI, [−0.32,0.54]; BF_10_=0.30). RDM-based results are not confounded by variability in signal strength or activation extent.

**Supplementary Figure S7.**
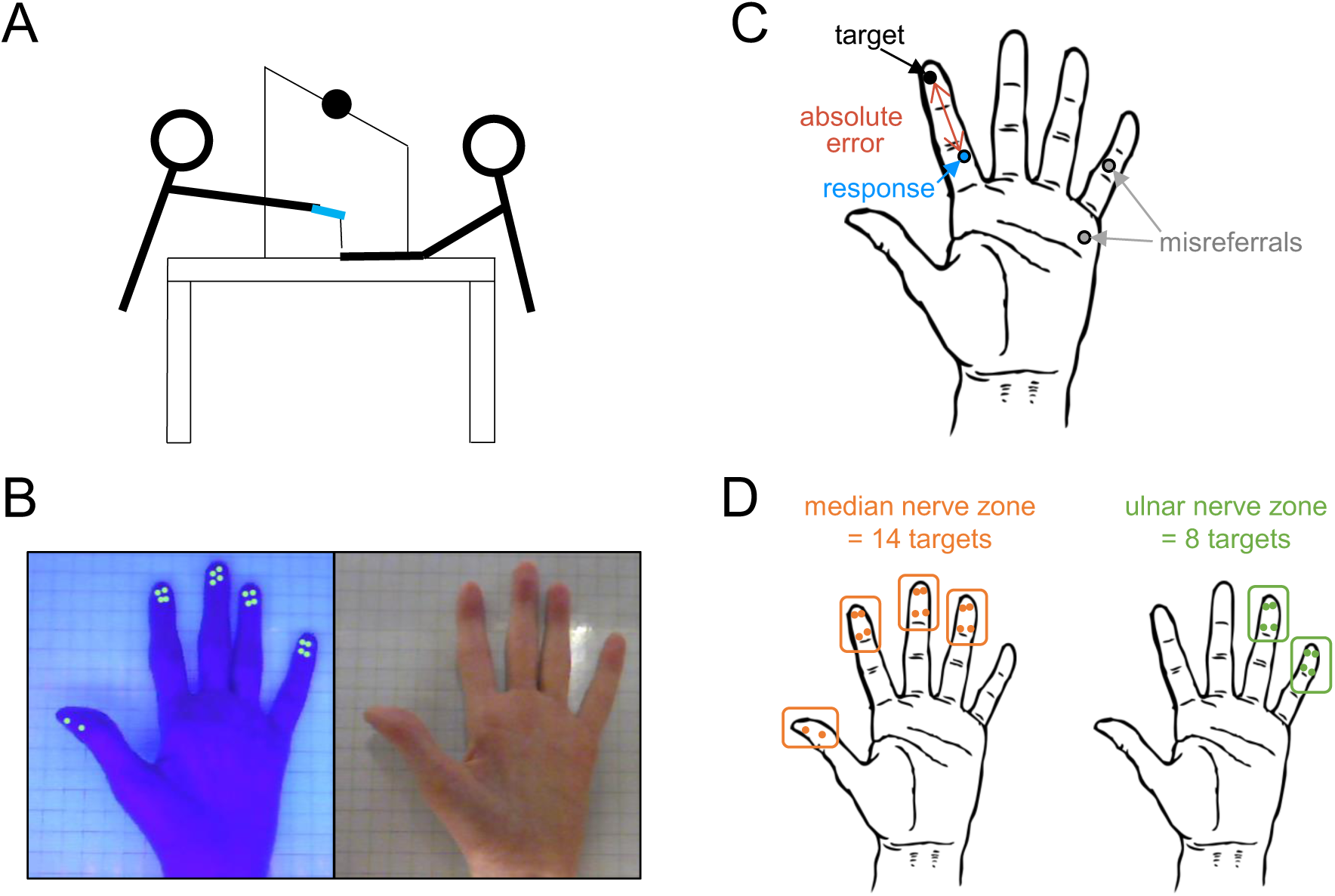
Locognosia testing: Digital photograph method. **A:** The experimenter (left) and participant (right) in position during testing. The participant’s hand is through a ‘blinder box’ to obscure their vision of the hand, and the experimenter applies stimulation using a monofilament. **B**: UV-light image (left) used by the experimenter to register targets and a normal light image (right) on which the participant registers their responses. **C**: The target is the location where the experimenter applies the touch stimulus. Response is where the participant indicates they felt they were touched. The absolute localization error is computed as the Euclidian distance between target and response. Responses made to another digit or to the palm are defined as misreferrals. **D**: Targets defined as within the repair zone for median (left, orange) and ulnar (right, green) nerve repairs. Part of this figure has been modified from Weber et al.^2^, licensed under CC-BY.

**Supplementary Figure S8.**
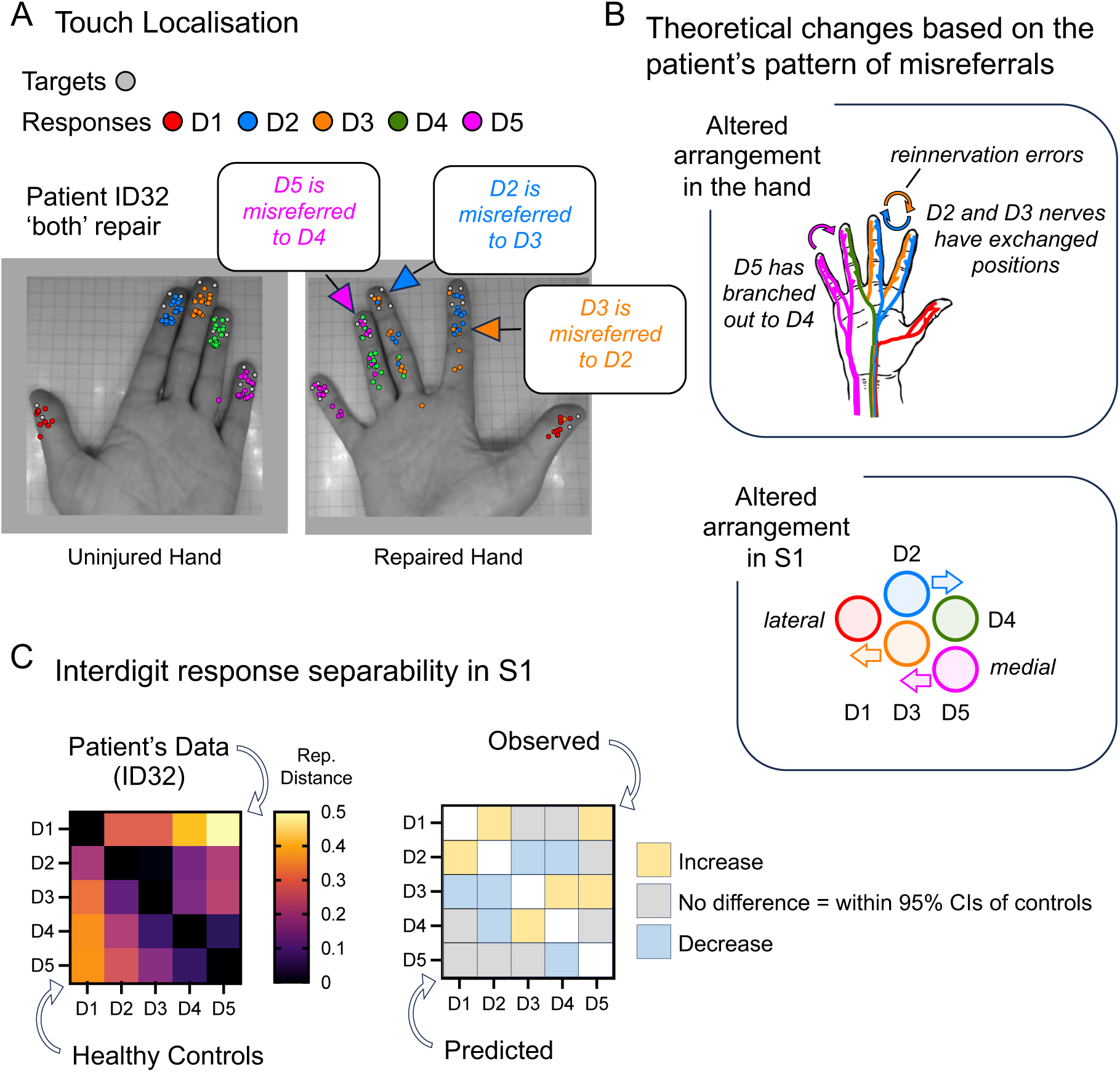
Frequent misreferrals between digits may reflect reinnervation errors: ID32. **A:** Locognosia results from patient ID32. Targets (grey circles) and responses (coloured circles) are overlaid on the photographs of the hand. The colours indicate which digits were touched (see inset key). Touch of D5 was frequently misreferred to D4, and D2 and D3 were often confused. **B:** Theoretical changes in the arrangement of nerves in the hand based on the patient’s pattern of misreferrals. Reinnervation errors may have resulted in a nerve of D5 re-routed to D4, and nerves of D2 and D3 may have exchanged. This would predictably alter the arrangement of digit maps in S1. For example, D5 should ‘move closer’ to D4. **C:** Representational distances are shown in the heatmap, with patient ID32’s data above the reference diagonal and the mean of healthy controls below (left). Predicted and observed differences between the patient’s data and controls (right). The observed differences are above the reference diagonal and predicted differences are below. For each interdigit comparison, ‘increased’/‘decreased’ observed results indicates that the patient’s representational distances are above/below the 95% confidence intervals of healthy controls, respectively.

**Supplementary Figure S9.**
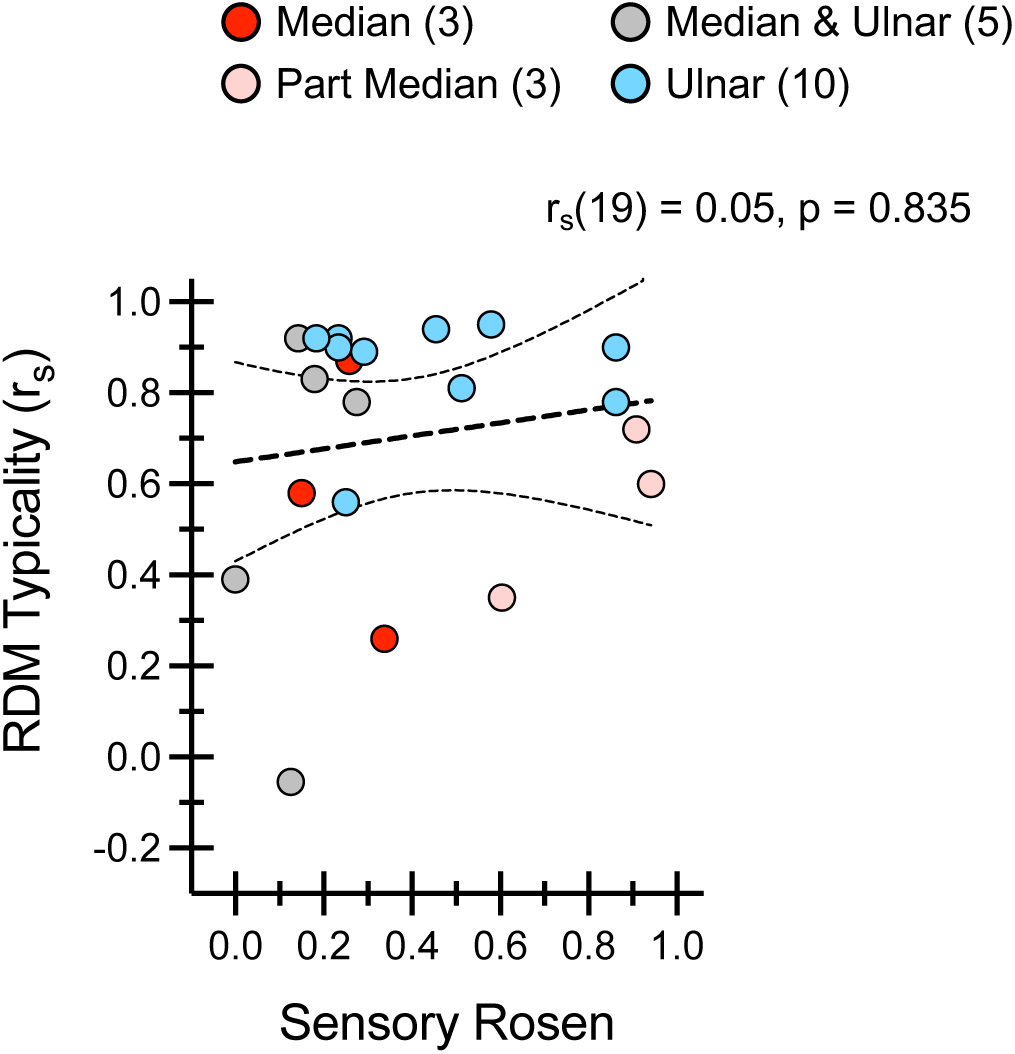
No evidence for a relationship between altered S1 maps and sensory Rosen scores. The sensory Rosen test measures both low- and high-level sensory and motor hand function (see Methods: *Sensory Rosen scores*). The best score possible is 1.0, indicating no impairment. Scatterplots show RDM typicality in *S1-patients-repaired* versus sensory Rosen scores. No evidence for a relationship is found (r_s_(19)=0.05, p=0.835; 95%-CI, [−0.40, 0.48]; BF_10_=0.35). ‘Part median’ indicates patients with incomplete median nerve transection injuries.

**Supplementary Figure S10.**
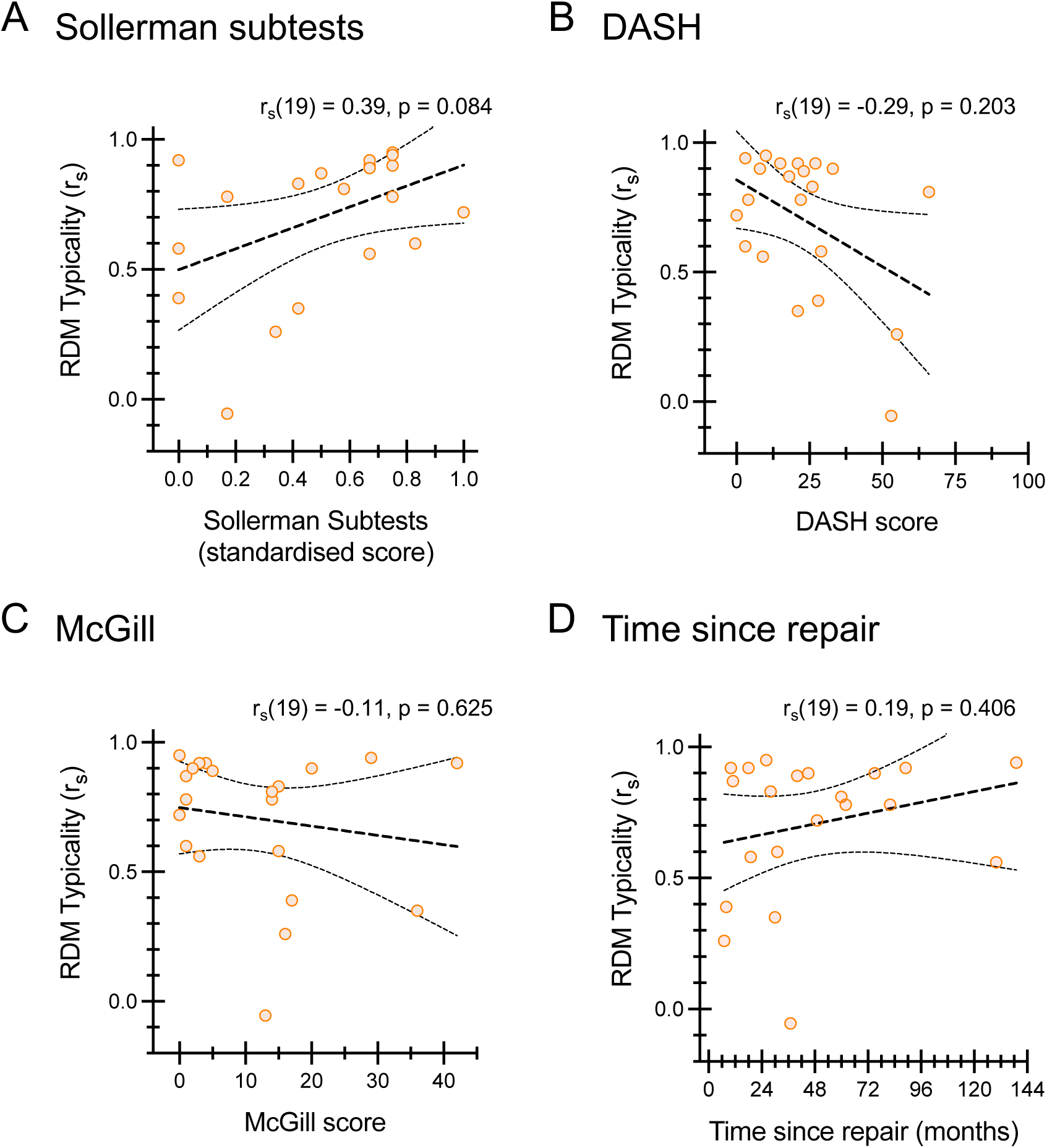
No evidence for a relationship between altered S1 maps and Sollerman subtest scores, DASH, McGill, or time since repair. Scatterplots show RDM typicality in *S1-patients-repaired* against Sollerman subtest scores (**A**), patient-reported disability, DASH scores (**B**), patient-reported pain, McGill scores (**C**), and time since repair (**D**). Across all analyses, we find no credible evidence for associations with RDM typicality (**A**: r_s_(19)=0.39, p=0.084; 95%-CI, [−0.06,0.71]; BF_10_=0.97 **B**: r_s_(19)=-0.29, p=0.203; 95%-CI, [−0.65,0.18]; BF_10_=0.80; **C**: r_s_(19)=-0.11, p=0.625; 95%-CI, [−0.53,0.35]; BF_10_=0.35; **D**: r_s_(19)=0.19, p=0.406; 95%-CI, [−0.27,0.59]; BF_10_=0.45. **A:** The Sollerman hand function subtests (see Methods: *Sensory Rosen scores*) evaluate goal-directed hand actions with relatively coarse metrics (completion time and subjective difficulty). A score of 1 indicates no impairment. **B:** The DASH (Disabilities of the Arm, Shoulder and Hand) is a general upper-limb disability measure (not specific to nerve injury). Because it is agnostic to how tasks are performed, compensatory strategies can mask deficits. A score of 100 indicates maximal difficulty. **C:** The short-form McGill questionnaire is a patient-reported outcome where scores reflect the severity of pain experienced. A score of 45 indicates maximal pain. **D:** Time since repair (from surgical repair to testing).

**Supplementary Figure S11.**
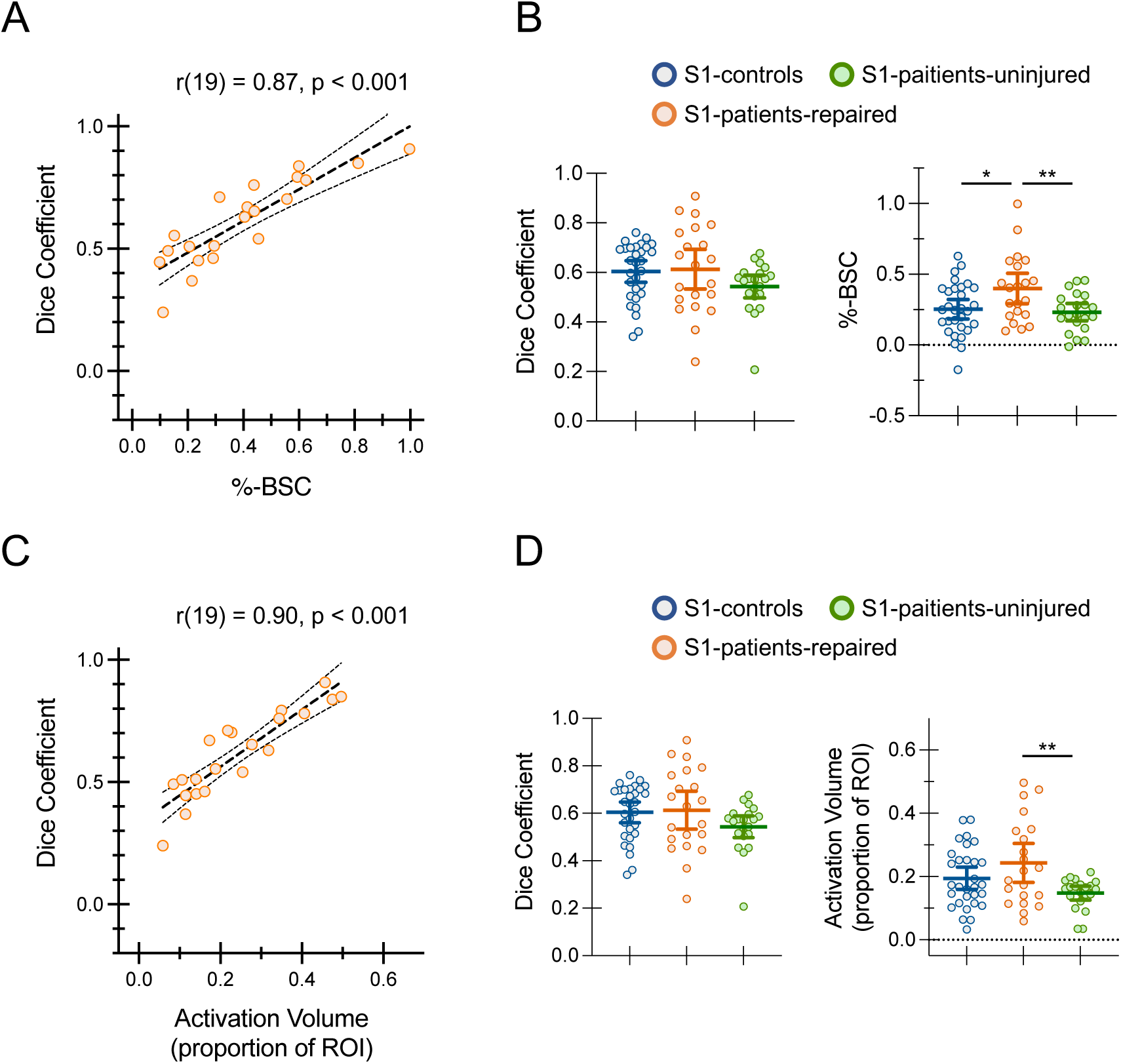
Dice coefficients correlate with %-BSC and activation volume. **A:** In *S1-patients-repaired*, Dice coefficients show a strong positive correlation with percent BOLD signal change (r(19) = 0.87, p < 0.001; 95%-CI, [0.70, 0.95]). **B:** Group-level values for *S1-patients-repaired*, *S1-patients-uninjured*, and *S1-controls* are plotted separately for Dice coefficients (left) and %-BSC (right), showing broadly similar between-group patterns. Statistical results—Dice: *S1-patients-repaired* vs *S1-controls* (unpaired t-test with Welch’s correction: t(32)=0.21, p=0.839; *η*^2^<0.01; 95%-CI, [−0.08,0.10]; BF_10_=0.29; *S1-patients-repaired* vs *S1-patients-uninjured* (t(20)=1.73, p=0.099; H–B p=0.198; *η*^2^=0.13; 95%-CI, [−0.01,0.15]; BF_10_=0.81); *S1-patients-uninjured* vs *S1-controls* (U=204, p=0.034; H–B p=0.101; r_rb_=-0.35; 95%-CI, [−0.12,-0.01]; BF_10_=0.32). Statistical results—%-BSC (same data as Fig. 6A of the main manuscript): *S1-patients-repaired* vs *S1-controls* (t(49)=2.47, p=0.017; H-B p=0.033; *η*^2^=0.11; 95%-CI, [0.03,0.27]); *S1-patients-repaired* vs *S1-patients-uninjured* (t(20)=3.65, p=0.002; H-B p=0.005; *η*^2^=0.40; 95%-CI, [0.07,0.26]); *S1-patients-uninjured* vs *S1-controls* (t(49)=0.45, p=0.657; *η*^2^<0.01; 95%-CI, [−0.12,0.07]; BF_10_=0.31). **C**: Dice coefficients also show a strong positive correlation with activation volume (r(19)=0.90, p < 0.001; 95%-CI, [0.76,0.96]). **D:** Same layout as in B, now showing group-level results for Dice coefficients (left) and activation volume (right). Statistical results—Activation volume: *S1-patients-repaired* vs *S1-controls* (t(49)=1.52, p=0.135; *η*^2^=0.04; 95%-CI, [−0.02,0.11]; BF_10_=0.72); *S1-patients-repaired* vs *S1-patients-uninjured* (t(20)=3.61, p=0.002; H–B p=0.005; *η*^2^=0.40; 95%-CI, [0.04,0.15]); *S1-patients-uninjured* vs *S1-controls* (unpaired t-test with Welch’s correction: t(45.5)=2.31, p=0.025; H–B p=0.051; *η*^2^=0.11; 95%-CI, [−0.09,-0.01]; BF_10_=1.61). Dice coefficients are tightly coupled with response magnitude and spatial extent, complicating their interpretation when comparing across groups. Error bars are 95% confidence intervals. Asterisks indicate multiple-comparison-corrected significance (* p < 0.05; ** p < 0.001; two-sided).

**Supplementary Figure S12.**
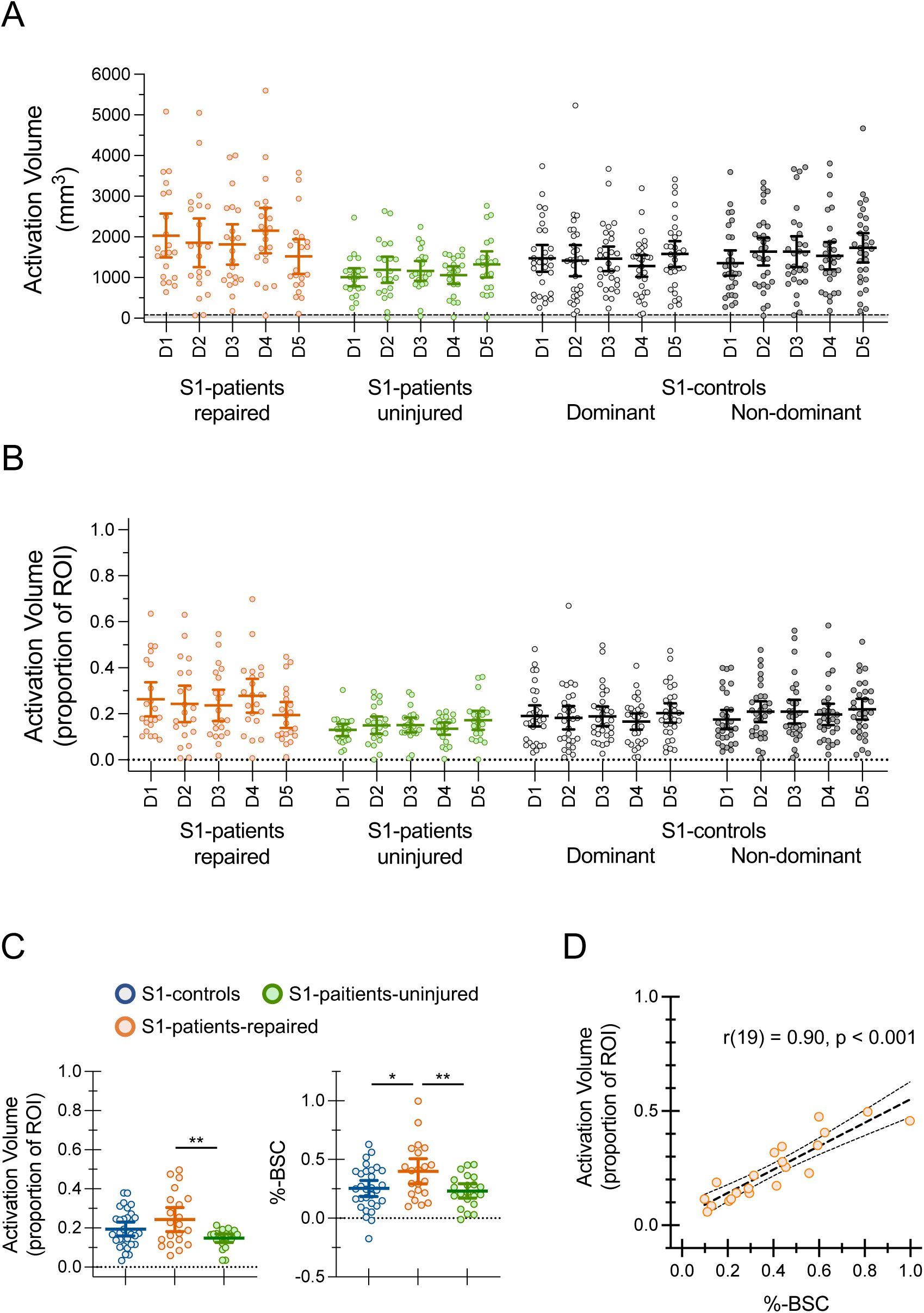
Activation volume. **A–B**: Activation volume per digit contrast (D1–D5) for S1-patients-repaired, *S1-patients-uninjured*, and *S1-controls* (shown separately for dominant and non-dominant hands). **A**: Data in 1mm^3^ voxels; 80mm^3^ (10 native functional voxels at 2mm^3^) is marked on the y-axis for reference. **B**: Same data expressed as a proportion of total ROI volume. While most participants show robust responses for all digits, instances of low activity occur in both patients and controls. **C:** Group results averaged across digits broadly mirror %-BSC differences. Statistical results—Activation volume: *S1-patients-repaired* vs *S1-controls* (t(49)=1.52, p=0.135; *η*^2^=0.04; 95%-CI, [−0.02, 0.11]; BF_10_=0.72); *S1-patients-repaired* vs *S1-patients-uninjured* (t(20)=3.61, p=0.002; *η*^2^=0.40; 95%-CI, [0.04, 0.15]); *S1-patients-uninjured* vs *S1-controls* (unpaired t-test with Welch’s correction: t(45.46)=2.31, p=0.025; H–B p=0.051; *η*^2^=0.11; 95%-CI, [−0.09, −0.01]; BF_10_=1.61]); Statistical results—%-BSC (same data as Fig. 6A of the main manuscript): *S1-patients-repaired* vs *S1-controls* (t(49)=2.47, p=0.017; H-B p=0.033; *η*^2^=0.11; 95%-CI, [0.03, 0.27]); *S1-patients-repaired* vs *S1-patients-uninjured* (t(20)=3.65, p=0.002; H-B p=0.005; *η*^2^=0.40; 95%-CI, [0.07, 0.26]); *S1-patients-uninjured* vs *S1-controls* (t(49)=0.45, p=0.657; *η*^2^<0.01; 95%-CI, [−0.12, 0.07]; BF_10_=0.31). **C:** Scatterplot showing activation volume versus %-BSC in *S1-patients-repaired* revealing a strong positive correlation (r(19)=0.90, p<0.001; 95%-CI, [0.76, 0.96]). Error bars are 95% confidence intervals. Asterisks indicate multiple-comparison-corrected significance (* p < 0.05; ** p < 0.01; two-sided).

Supplementary Table S1 provides patient demographics, standardised clinical assessment scores, and locognosia performance for their repaired hand.

**Supplementary Table S1.** Patient demographics, standardised clinical assessments, and locognosia performance.

| Subject ID | Demographics |  |  |  |  | Standardised Clinical Assessments |  |  |  | Locognosia<sup>f</sup> |  |
| --- | --- | --- | --- | --- | --- | --- | --- | --- | --- | --- | --- |
|  | Sex | WS<sup>a</sup> | MSR | Side | Nerve | DASH<sup>b</sup> | McGill<sup>c</sup> | Rosen |  | Error | Mis. |
|  |  |  |  |  |  |  |  | Sensory<sup>d</sup> | Pain<sup>e</sup> |  |  |
| 21 | F | 28 | 19 | L | M | 29 | 15 | 0.15 | 0.17 | 11.3 | 0.40 |
| 22 | M | 30 | 10 | L | U | 21 | 3 | 0.23 | 0.33 | 16.5 | 0.02 |
| 25 | M | 30 | 60 | R | U | 66 | 14 | 0.51 | 0 | 10.3 | 0.04 |
| 26 | F | 30 | 37 | R | M+U | 53 | 13 | 0.13 | -- | 14.3 | 0.13 |
| 27 | F | -8 | 62 | R | U | 4 | 1 | 0.86 | 0.67 | 4.1 | 0.13 |
| 30 | F | 30 | 26 | L | U | 10 | 0 | 0.58 | 0.67 | 4.3 | 0.00 |
| 31 | M | 15 | 75 | R | U | 33 | 20 | 0.86 | 0.67 | 6.3 | 0.00 |
| 32 | M | 30 | 82 | R | M+U | 22 | 14 | 0.28 | 0.5 | 19.5 | 0.27 |
| 33 | F | 29 | 11 | L | M | 18 | 1 | 0.26 | 0.5 | 10.6 | 0.06 |
| 34 | F | 30 | 8 | L | M+U | 28 | 17 | 0.00 | 0 | 22.6 | 0.50 |
| 35 | M | 30 | 28 | L | M+U | 26 | 15 | 0.18 | 0.5 | 16.2 | 0.07 |
| 36 | M | 30 | 139 | R | U | 3 | 29 | 0.0 | 0.67 | -- | -- |
| 38 | M | 30 | 18 | R | M+U | 15 | 4 | 0.14 | 0.5 | 16.6 | 0.07 |
| 39 | M | 30 | 30 | L | M<sup>g</sup> | 21 | 36 | 0.60 | 1.0 | 8.4 | 0.01 |
| 40 | M | 22 | 31 | R | M<sup>g</sup> | 3 | 1 | 0.94 | 0.83 | 4.8 | 0.01 |
| 42 | F | 27 | 45 | R | U | 8 | 2 | 0.23 | 0.67 | 5.8 | 0.02 |
| 46 | M | 30 | 49 | L | M<sup>g</sup> | 0 | 0 | 0.91 | 1.0 | 8.3 | 0.00 |
| 47 | M | -29 | 89 | L | U | 27 | 42 | 0.18 | 0.33 | -- | -- |
| 57 | M | -30 | 130 | R | U | 9 | 3 | 0.25 | 0.67 | 12.0 | 0.14 |
| 61 | M | 3 | 40 | L | U | 23 | 5 | 0.29 | 0.67 | 9.5 | 0.03 |
| 63 | M | 30 | 7 | R | M | 55 | 16 | 0.34 | 0.33 | 32.8 | 0.18 |

Sex: F = female, M = male; WS = Waterloo score; MSR = months since repair; Side: L = left, R = right; Nerve: M = median, U = ulnar, M+U = combined median and ulnar repairs; Mis. = Misreferrals. -- indicates missing values.
<sup>a</sup> Modified Waterloo Handedness Inventory<sup>1</sup> is a questionnaire used to evaluate hand preference. Scores range from -30 to 30, where negative values indicate left-hand preference, and positive values indicate right-hand preference.
<sup>b</sup> The Disabilities of the Arm, Shoulder and Hand (DASH)<sup>2</sup> is a patient-reported outcome questionnaire where scores indicate the level of difficulty experienced using the upper limb to perform activities of daily living. A score of 0 indicates no difficulty; a score of 100 indicates maximal difficulty. This is a general test of upper limb function, not specific to nerve injury.
<sup>c</sup> The short-form McGill<sup>3</sup> questionnaire is a patient-reported outcome where scores reflect the severity of pain experienced. A score of 0 indicates no pain; a score of 45 indicates maximal pain.
<sup>d</sup> The sensory Rosen<sup>4</sup> test produces a single composite score, with subtests measuring touch detection thresholds using Semmes-Weinstein monofilaments, two-point discrimination, shape-texture-identification<sup>5</sup>, and the Sollerman<sup>6</sup> hand function subtests 4, 8, and 10. Higher scores indicate better function, with a score of 1 indicating no impairment. A score of 0 indicates maximal impairment (i.e., no sub-tests were completed successfully).
<sup>e</sup> The Pain domain of the Rosen test is a patient-reported outcome questionnaire that evaluates pain and discomfort due to nerve injury. Higher scores indicate less pain/discomfort, with a score of 1 indicating no/minor pain/discomfort. A score of 0 indicates maximal pain/discomfort (i.e., "hinders function").
<sup>f</sup> Locognosia was evaluated using the Digital Photograph Method<sup>7</sup>. Error is the absolute error of localisation in mm, excluding misreferrals. Misreferrals are errors made between digits or from a digit to the palm, expressed in the table as proportions (i.e., the total number of misreferrals divided by the total number of stimulation trials). Notably, data were previously reported in Weber et al.<sup>7</sup> using different subject IDs, labelled as "P", but the participant order presented here remains unchanged: ID21 = P1, ID22 = P2, and so on, up to ID61 = P18. ID63 was tested more recently.
<sup>g</sup> Patients with incomplete nerve transection injuries.

## Supplementary Table S2. Dice results

This section summarises the results of the Dice analysis.

## Methods

Five contrasts were performed per hand per participant: D1 > rest, D2 > rest, D3 > rest, D4 > rest, D5 > rest. Resultant activity was thresholded at z = 2.0, uncorrected, selecting only positive-going responses. Only active voxels within the anatomically defined contralateral S1 ROI were used to compute dice coefficients, quantifying the overlap between digit response maps. Dice coefficients were computed using the minimum-cluster normalised method, dividing the number of shared voxels by the number of voxels in the smaller map. If a smaller map sits entirely within a larger map, the dice coefficient is 1.

## Interpretational caveat

As detailed in Methods (*Measuring digit map separability*), Dice values are strongly influenced by response robustness. They correlate closely with both %-BSC (r = 0.87, p < 0.001; Supplementary Fig. S11A) and activation volume (r = 0.90, p < 0.001; Supplementary Fig. S11B). Group differences in Dice may therefore reflect differences in signal strength rather than true differences in map separability. Results are provided here for completeness and transparency.

**Table S2A.** Testing for differences between hands in healthy controls.

| Independent Variable | Dependent Variable | Test | Result |
| --- | --- | --- | --- |
| hand | Mean dice | Wilcoxon signed rank test | $W = 238$, $p = .919$, $r_{rb} = -0.02$, 95% CI = [-0.033, 0.037] |
| repair zone | Mean dice | paired t-test | $t(29) = -1.22$, $p = .233$, $\eta^2 = 0.05$, 95% CI = [-0.072, 0.018] |
| hand | Dice typicality | Wilcoxon signed rank test | $W = 191$, $p = .574$, $r_{rb} = -0.05$, 95% CI = [-0.155, 0.075] |

This table shows the results of testing for differences between the two hands of healthy controls. Hand = data represents the entire hand, testing for differences between dominant and non-dominant hands. Repair-zone = healthy controls were assigned a simulated injured condition matched to patients (see Methods); data represents the simulated repair zone, testing for differences between hands. 95% CI = 95% confidence interval.

**Table S2B.**
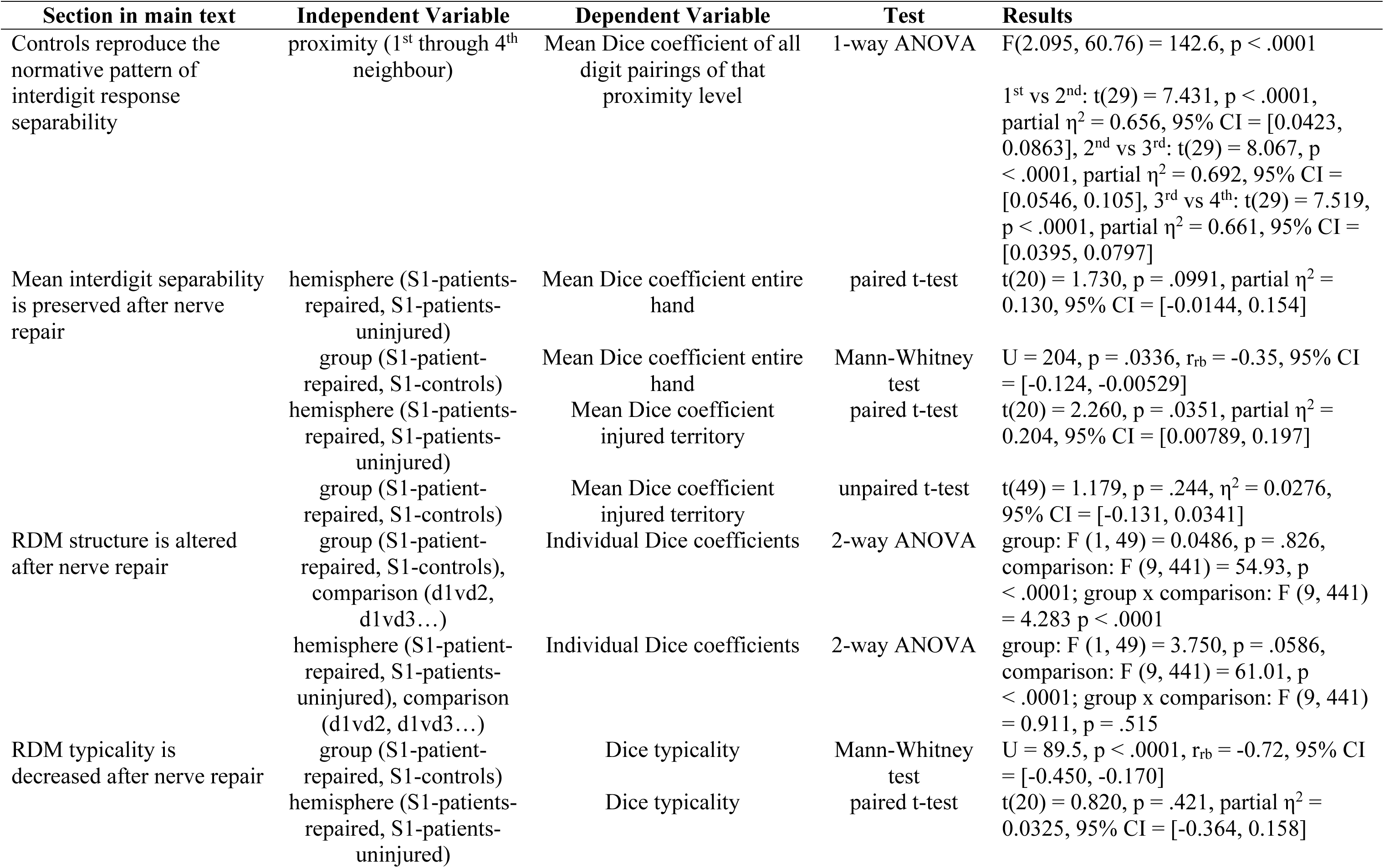
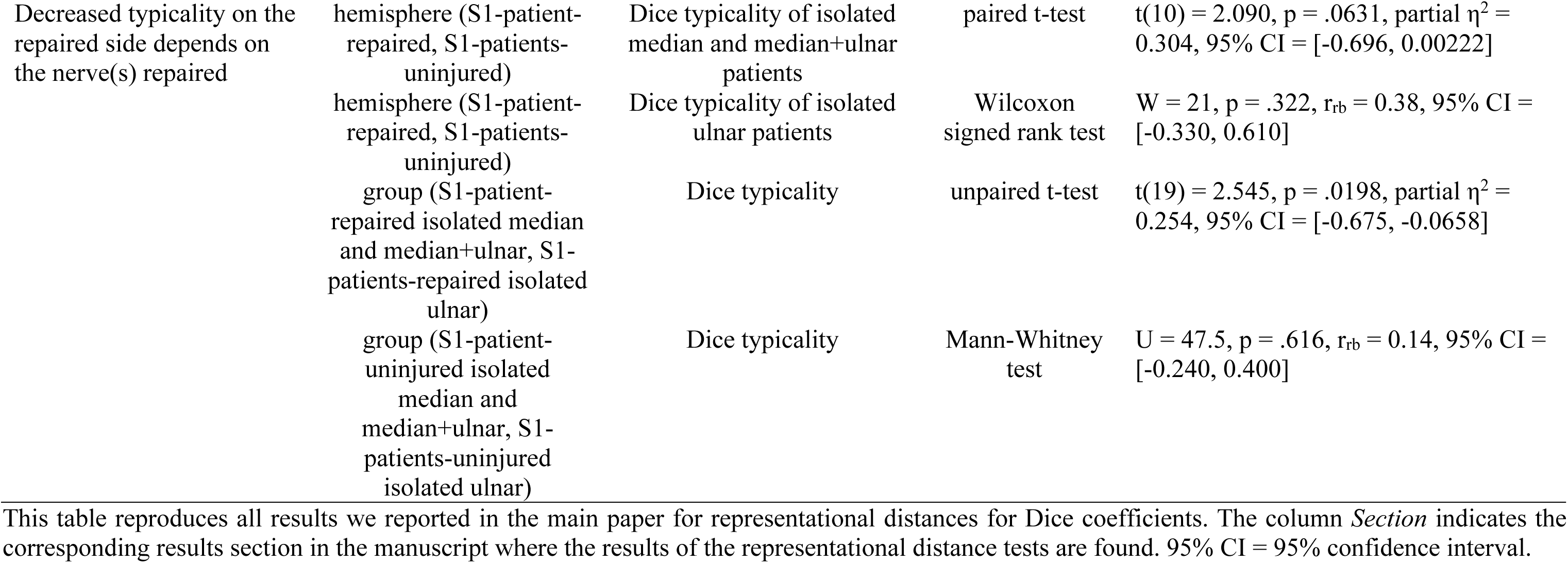
Dice results for all main-text analyses.

**Supplementary Table S3.** Testing for differences between hands in healthy controls.

| Independent Variable | Dependent Variable | Test | Result |
| --- | --- | --- | --- |
| hand | mean rep. distance | Wilcoxon signed rank test | $W = 227, p = .919, r_{rb} = -0.02, 95\% \text{ CI} = [-0.042, 0.033]$ |
| repair zone | mean RDM | Wilcoxon signed rank test | $W = 262, p = .556, r_{rb} = 0.13, 95\% \text{ CI} = [-0.021, 0.039]$ |
| hand | typicality | Wilcoxon signed rank test | $W = 257, p = .621, r_{rb} = -0.11, 95\% \text{ CI} = [-0.02, 0.04]$ |
| hand | mean RDM reliability | Paired t-test | $t(29) = 0.612, p = .545, \eta^2 = 0.01, 95\% \text{ CI} = [-0.043, 0.079]$ |
| hand | mean %BSC | Paired t-test | $t(29) = -1.67, p = .106, \eta^2 = 0.01, 95\% \text{ CI} = [-0.12, 0.012]$ |
| repair zone | mean %BSC | Wilcoxon signed rank test | $W = 205, p = .584, r_{rb} = -0.06, 95\% \text{ CI} = [-0.109, 0.063]$ |

This table shows the results of testing for differences between the two hands of healthy controls for several measures. Hand = data represents the entire hand, testing for differences between dominant and non-dominant hands. Repair-zone = healthy controls were assigned a simulated injured condition matched to patients (see Methods); data represents the simulated repair zone, testing for differences between hands. 95% CI = 95% confidence interval.

**Supplementary Table S4.** Sensory Rosen constituent component scores.

| Subject ID | SWM raw | SWM norm. | 2PD raw | 2PD norm. | STI raw | STI norm. | Sollerm. raw | Sollerm. norm. | Sensory score |
| --- | --- | --- | --- | --- | --- | --- | --- | --- | --- |
| 21 | 4.00 | 0.27 | 1.00 | 0.33 | 0.00 | 0.00 | 0.00 | 0.00 | 0.15 |
| 22 | 4.00 | 0.27 | 0.00 | 0.00 | 0.00 | 0.00 | 8.00 | 0.67 | 0.23 |
| 25 | 7.00 | 0.47 | 0.00 | 0.00 | 6.00 | 1.00 | 7.00 | 0.58 | 0.51 |
| 26 | 5.00 | 0.33 | 0.00 | 0.00 | 0.00 | 0.00 | 2.00 | 0.17 | 0.13 |
| 27 | 13.00 | 0.87 | 3.00 | 1.00 | 5.00 | 0.83 | 9.00 | 0.75 | 0.86 |
| 30 | 13.50 | 0.90 | 0.50 | 0.17 | 3.00 | 0.50 | 9.00 | 0.75 | 0.58 |
| 31 | 13.00 | 0.87 | 3.00 | 1.00 | 5.00 | 0.83 | 9.00 | 0.75 | 0.86 |
| 32 | 9.00 | 0.60 | 0.00 | 0.00 | 2.00 | 0.33 | 2.00 | 0.17 | 0.28 |
| 33 | 8.00 | 0.53 | 0.00 | 0.00 | 0.00 | 0.00 | 6.00 | 0.50 | 0.26 |
| 34 | 0.00 | 0.00 | 0.00 | 0.00 | 0.00 | 0.00 | 0.00 | 0.00 | 0.00 |
| 35 | 4.50 | 0.30 | 0.00 | 0.00 | 0.00 | 0.00 | 5.00 | 0.42 | 0.18 |
| 36 | 8.50 | 0.57 | 1.50 | 0.50 | 0.00 | 0.00 | 9.00 | 0.75 | 0.45 |
| 38 | 8.50 | 0.57 | 0.00 | 0.00 | 0.00 | 0.00 | 0.00 | 0.00 | 0.14 |
| 39 | 10.00 | 0.67 | 2.00 | 0.67 | 4.00 | 0.67 | 5.00 | 0.42 | 0.60 |
| 40 | 14.00 | 0.93 | 3.00 | 1.00 | 6.00 | 1.00 | 10.00 | 0.83 | 0.94 |
| 42 | 4.00 | 0.27 | 0.00 | 0.00 | 0.00 | 0.00 | 8.00 | 0.67 | 0.23 |
| 46 | 12.00 | 0.80 | 3.00 | 1.00 | 5.00 | 0.83 | 12.00 | 1.00 | 0.91 |
| 47 | 1.00 | 0.07 | 0.00 | 0.00 | 0.00 | 0.00 | 8.00 | 0.67 | 0.18 |
| 57 | 5.00 | 0.33 | 0.00 | 0.00 | 0.00 | 0.00 | 8.00 | 0.67 | 0.25 |
| 61 | 5.00 | 0.33 | 0.00 | 0.00 | 0.00 | 0.00 | 8.00 | 0.67 | 0.25 |
| 63 | 9.00 | 0.60 | 1.00 | 0.33 | 0.00 | 0.00 | 5.00 | 0.42 | 0.34 |

The Sensory Rosen score is derived from four component tests: Semmes–Weinstein monofilaments (SWM), two-point discrimination (2PD), shape and texture identification (STI), and the sensory subtests of the Sollerman hand function test (Sollerm.). Each test yields a raw point score, which is then converted to a normalised score by dividing the raw score by the normative maximum score from healthy controls (Hakir. National Assessment Manual: For assessment of hand function after nerve repair. 2018. https://hakir.se/about-hakir/). The overall Sensory Rosen score is the mean of the normalised component scores. This table reports raw and normalised scores for each test, as well as the final composite score. Abbreviations: SWM = Semmes–Weinstein monofilaments; 2PD = two-point discrimination; STI = shape and texture identification; Sollerm. = Sollerman; norm. = normalised.

